# Predicting Substance Use and Psychotic-Like Experiences in Adolescents

**DOI:** 10.64898/2026.05.20.26353709

**Authors:** Carolyn M. Amir, Catherine Walsh, Haley R. Wang, Dara G. Ghahremani, Sarah E. Chang, Tiffany C. Ho, Lucina Q. Uddin, Ziva D. Cooper, Jesse Rissman, Carrie E. Bearden

**Author notes:** Correspondence should be addressed to: Carrie E. Bearden, Ph.D., A7-460 Semel Institute, Los Angeles, CA 90095.

## Abstract

Adolescence is a critical developmental window for the emergence of substance use and psychosis-spectrum symptoms, yet early risk for these outcomes remains poorly understood. Using longitudinal data from the Adolescent Brain Cognitive Development (ABCD) Study (n=10,134), we tested whether demographic, clinical, and structural and functional neuroimaging measures assessed in childhood (mean baseline age=9.96 years) predict later adolescent substance use, psychotic-like experiences, and/or their co-occurrence. Multivariate machine learning models reliably predicted later emergence of psychotic-like experiences (AUROC=0.780) and their co-occurrence with substance use (AUROC= 0.828), as well as substance use on its own (AUROC=0.626). Distinct patterns of functional brain connectivity, task-related brain activation, demographic, and clinical factors differentiated each outcome. Findings suggest that partially dissociable developmental risk profiles are detectable as early as childhood, and results underscore the importance of explicitly modeling comorbidity when interrogating risk factors for mental health outcomes.

---

There is a critical need to understand how demographic, environmental, clinical, and neural factors jointly shape individual risk for psychiatric illness. Predicting risk before the onset of overt disorder is an important clinical goal with substantial translational potential. When applied to longitudinal data, classification-based machine learning approaches offer a powerful framework for predicting adverse mental health outcomes before they manifest.^1–4^ Improving the sensitivity of such models to detect early risk is essential for enabling timely, targeted interventions.^4–6^ At the same time, advances in analytic tools, including gradient-boosted machine learning models, allow for more precise characterization of neurobiological risk profiles at the group level.^7,8^ Here, we integrated advanced machine learning methods with a rich, longitudinal neuroimaging dataset incorporating multimodal imaging methods, thereby supporting a robust approach for predicting psychiatric illness risk in youth and deepening our understanding of the complex interplay between early risk factors.

Substance use (SU) is closely linked to risk for psychosis, particularly during child and adolescent development,^9,10^ making the identification of early markers of psychopathology a major public health priority. This need is amplified by evidence that substance exposure has age-dependent effects on the developing brain,^11,12^ alongside increasingly permissive drug legislation,^13^ and the growing popularity of vaping.^14–17^ Despite this growing need, identifying such risk factors remains challenging. SU and psychotic disorders show high comorbidity,^18^ and accumulating evidence indicates that adolescents displaying subthreshold psychotic symptoms are more likely to engage in SU.^19,20^

Early markers of vulnerability may be observable as early as childhood. Subthreshold psychotic-like experiences (PLEs) are relatively common during childhood and early adolescence^21,22^ and are associated with functional impairment and elevated risk for later psychopathology.^23,24^ Additionally, even low-level SU puts youth at higher risk for later SU disorders in adulthood.^25–28^

During child and adolescent development, the brain undergoes significant structural and functional maturation, making it particularly sensitive to environmental influences.^29–32^ Neurobiological changes during this critical window are governed by interactive processes influencing adolescent brain development - for instance, the family environment, socioeconomic status, and early adversity (for a recent review, see 29). Identifying early predictors from both neural and non-neural domains may clarify how the interplay of complex risk factors during a period of heightened plasticity jointly confers risk.

Substances commonly used during adolescence engage neural circuits that also play a central role in psychosis-spectrum symptoms, including frontostriatal and limbic circuitry implicated in reward processing,^21,33,34^ salience processing,^34–36^ and frontoparietal control networks.^37^ This overlap suggests that vulnerability to adolescent SU and psychotic spectrum symptoms may arise from shared, transdiagnostic risk processes, rather than substance-specific effects. Such convergence complicates efforts to disentangle substance-related effects from broader liability for psychosis. Consequently, a critical gap in the literature is the identification of factors that differentially confer risk for psychosis-spectrum symptoms in the absence of SU, SU in the absence of psychosis-spectrum symptoms, and their co-occurrence.

This study leverages the translational potential of multimodal data from the Adolescent Brain Cognitive Development (ABCD) Study by using a theoretically-motivated set of childhood risk factors to predict risk for SU, psychosis spectrum, and co-occurring outcomes in adolescence via machine learning classifiers and multivariate neuroimaging approaches. Multiple supervised classification algorithms were evaluated, consistent with our preregistered analysis plan. We hypothesized that: 1) multivariate machine learning models incorporating childhood neuroimaging, clinical, and demographic features would predict adolescent psychosis-spectrum symptoms, SU, and their co-occurrence; 2) each outcome would be characterized by a distinct combination of predictors, with both overlapping and outcome-specific features; and 3) co-occurring outcomes would reflect a partially distinct risk signature with shared and unique predictors relative to each single-outcome model.

## MAIN

## RESULTS

### 1. Models accurately predict psychotic-like experiences, substance use, and co-occurrence outcomes

Across all classification tasks, each of which involved distinguishing outcome-positive participants (e.g., adolescents with PLE) from outcome-negative participants (those with no PLE or SU outcome; ON), multivariate machine learning models trained on baseline data demonstrated consistent above-chance predictive performance. XGBoost models achieved the highest mean classification performance in 10 of 18 analyses, including 5 of 6 neuroimaging-only models (Supplementary Table 2b). Subsequent analyses therefore focus on XGBoost. Models were highly stable and achieved good performance for psychotic-like experiences (XGBoost mean area under the receiver operating curve [AUROC]=0.780, SD = 0.050, range = 0.722–0.826) and for co-occurrence of both outcomes (XGBoost mean AUROC=0.828, SD = 0.023, range = 0.803–0.863, and predicted SU at comparable accuracy the prior ABCD-based prediction study of early adolescent substance initiation (XGBoost mean AUROC=0.626; SD = 0.007, range across outer splits = 0.616–0.633) (Fig. 1A; ROC curves shown for best-performing split; full XGBoost performance metrics including sensitivity, specificity, and calibration in Supplementary Table 2a).^38^

**Figure 1.**
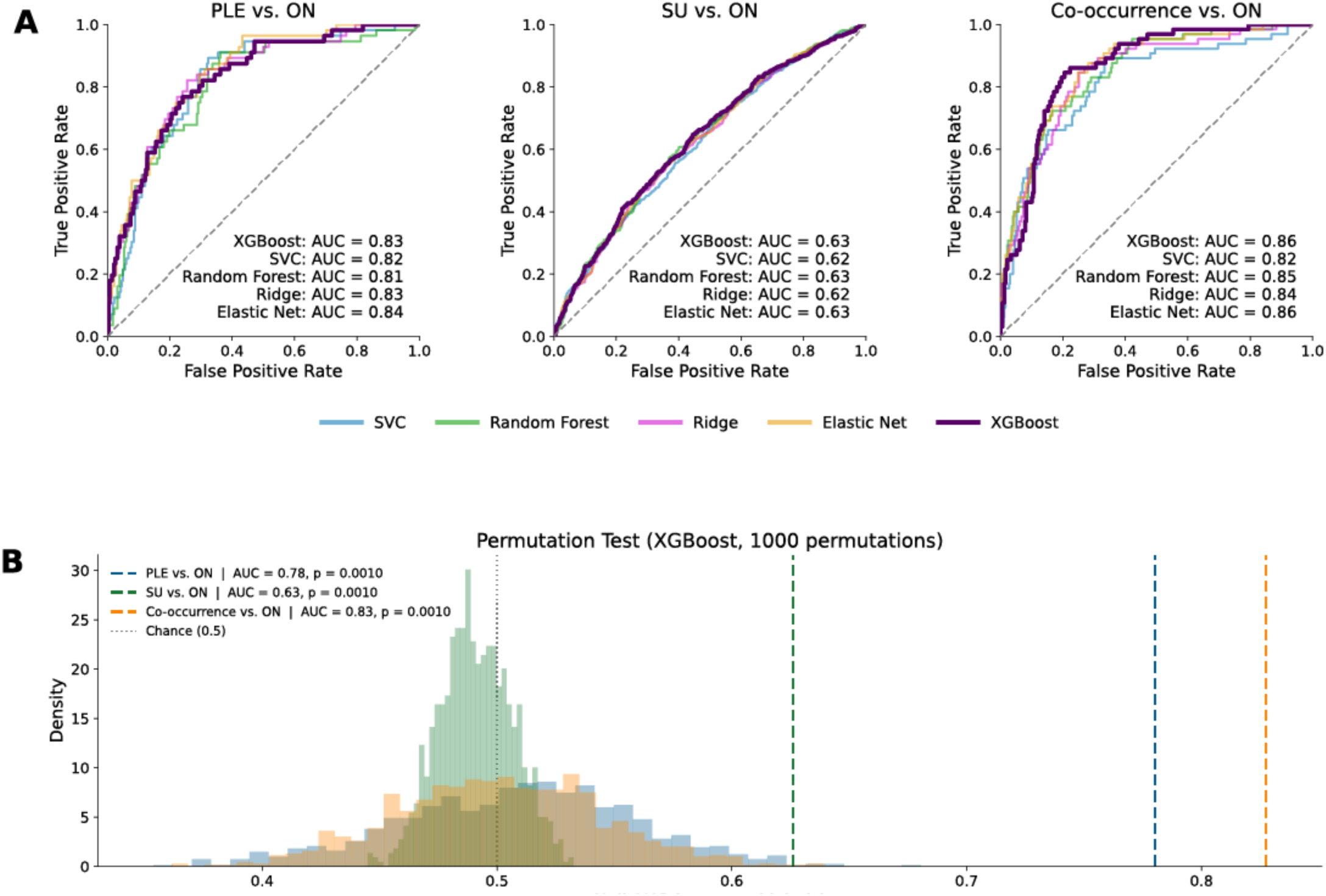
A) Receiver operating characteristic (ROC) curves for three binary prediction tasks: PLEs versus outcome-negative (ON; left), SU versus ON (middle), and co-occurring PLEs and SU versus ON (right), shown for the single best-performing outer cross-validation split for illustrative purposes only (mean AUROC across all splits reported in Supplementary Table 2). Models were trained on baseline demographic, clinical, and multimodal neuroimaging features and evaluated on held-out test data. Curves show true positive rate as a function of false positive rate across decision thresholds; the dashed diagonal indicates chance performance. Shown are Support Vector Classifier (SVC), Random Forest, Ridge, Elastic Net, and XGBoost models, with AUROC values for the best-performing split reported in each panel. Non-linear models, particularly XGBoost, showed the strongest discrimination across outcomes, with the largest gains for SU and co-occurrence. XGBoost results are therefore reported in subsequent analyses. B) Permutation test null distributions (1,000 permutations) for XGBoost across all three primary comparisons. Dashed vertical lines represent a chance line at AUC = .50; all three significantly exceeded the null distribution (all *p*<0.001). Abbreviations: PLE = psychotic-like experiences outcome group; SU = substance use outcome group; ON = outcome-negative group

Predictive performance consistently exceeded null models under label permutation (Figure 1B), indicating that accuracy depended on the correct alignment between features and outcomes. Model performance was stable across cross-validation folds and random seeds (Supplementary Figure 2). Consistent with these findings, XGBoost performance for each primary comparison significantly exceeded chance under permutation testing (Fig. 1B; 1,000 permutations; all *p* < 0.001). Precision-recall curves for primary comparisons are provided in Supplementary Figure 3, and calibration curves in Supplementary Figure 4. Negative control analyses using randomly permuted labels similarly confirmed a collapse to chance performance when true outcome structure was removed (Supplementary Table 3).

### 2. Distinct neuroimaging, clinical, and demographic features at baseline predict onset of psychosis spectrum symptoms and substance use endorsement in adolescence

The Shapley additive explanations (SHAP) derived from the XGBoost model identified baseline PLEs as the strongest predictor of subsequent PLEs, followed by fluid cognition, behavioral inhibition system (BIS) sensitivity, and sex (Fig. 2; Supplementary Figure 5), where positive feature-SHAP correlations indicate risk-increasing effects and negative correlations indicate protective effects. Sensitivity analyses revealed that differences were modest (mean |Δ| = 0.041) when baseline PLEs were dropped from the model entirely, indicating that the predictive signal in our primary models was not driven by baseline PLE inclusion. (Supplementary results, section 7). Additional demographic, environmental and clinical risk factors, including lower income, lower parental education, higher positive urgency, higher adverse childhood experiences (ACEs), and family history of depression also contributed substantially to PLE risk. Among neuroimaging features, lower right frontoparietal network (FPN)-accumbens resting state functional connectivity (rsFC), lower default mode network (DMN) within-network connectivity, lower forceps minor total diffusion (TD), and higher IFG pars triangularis transverse diffusivity were among the most influential predictors. Higher IFG activation during response inhibition (SST: inhibition vs. baseline and inhibition vs. go) showed modest protective effects.

**Figure 2.**
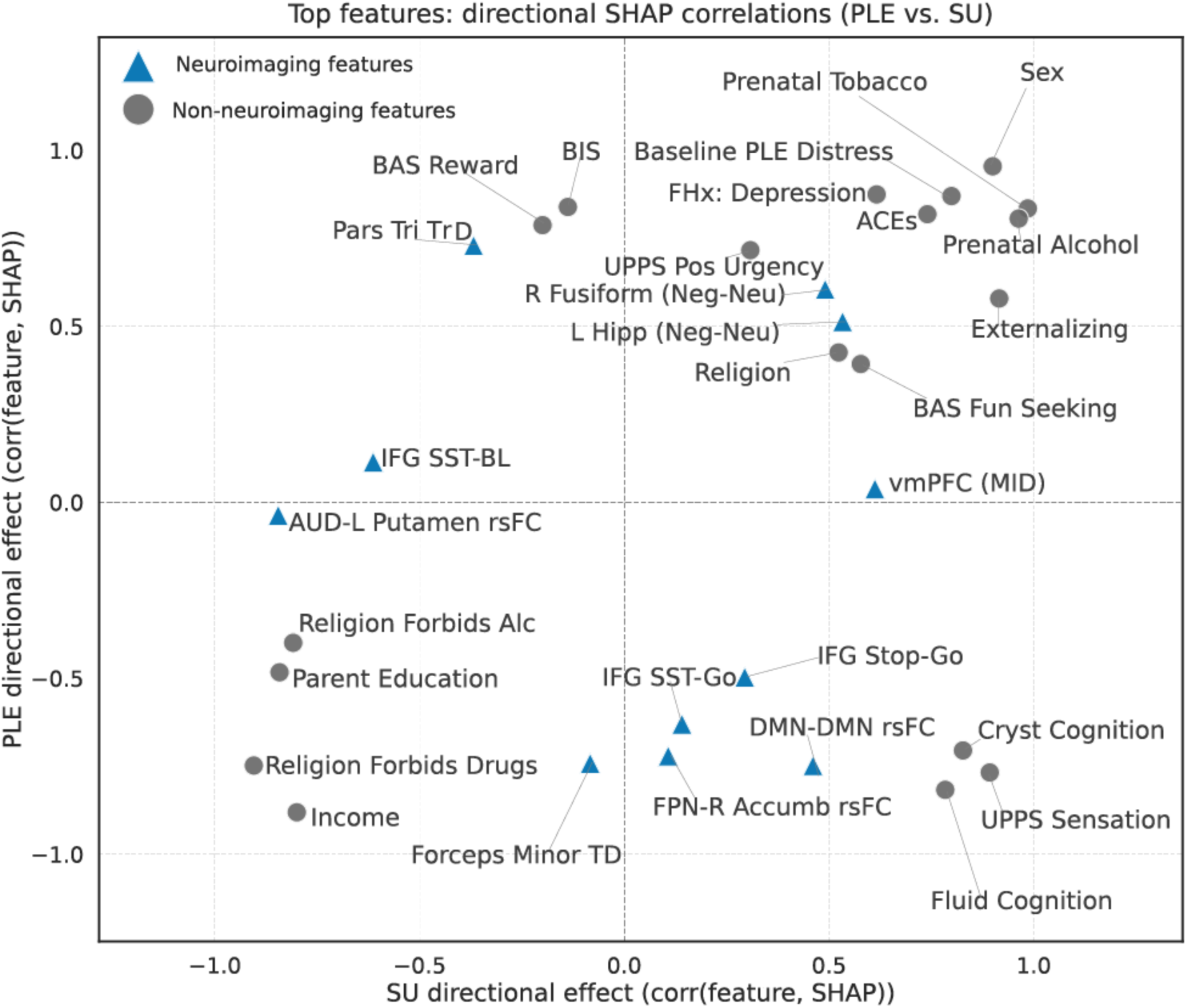
Directional feature importance for psychotic-like experiences (PLEs) and substance use (SU) derived from XGBoost SHAP analyses. A) Scatterplot showing directional associations between each feature and its SHAP value for SU (x-axis) and PLEs (y-axis), computed as the sign of the correlation between feature values and SHAP contributions on held-out test data, aggregated across cross-validation splits (see Supplementary Methods, Section 8). Displayed features comprise the union of the top 30 features ranked by combined PLE and SU mean absolute SHAP value and the top features identified across the three primary comparisons, ensuring that features contributing substantially to any single-outcome or co-occurrence model are represented. Each point represents a feature; triangles denote neuroimaging features and circles denote non-neuroimaging features. Distance from the origin reflects the strength of directional influence, with positive values indicating risk-increasing associations and negative values indicating protective associations. Dashed lines denote zero correlation, separating concordant and divergent effects across outcomes. Sex at birth was coded as binary; positive SHAP values indicate higher risk associated with female sex. **Abbreviations:** ACEs = Adverse Childhood Experiences; BIS = Behavioral Inhibition System; UPPS = UPPS Impulsive Behavior Scale; IFG = inferior frontal gyrus; Pars Tri TrD = Transverse diffusivity (grey matter) of the IFG pars triangularis; DMN = Default Mode Network; rsFC = resting-state functional connectivity; Cryst cognition = crystallized cognition; SST = Stop Signal Task; IFG SST-Go = IFG activation during response inhibition (SST: inhibition vs. baseline); IFG Stop-go: IFG activation during response inhibition (SST: inhibition vs. go); IFG SST-BL = IFG activation during correct-go response execution (SST: correct-go vs. fixation); vmPFC = ventromedial prefrontal cortex; MID = Monetary Incentive Delay Task; BAS = Behavioral Activation System; AUD = Auditory network; Accumb = nucleus accumbens; FPN = frontoparietal network; R fusiform = right fusiform gyrus; Neg-Neu = the negative faces versus neutral faces contrast from the Emotional nBack Task; Forceps Minor TD = restricted total diffusion (white matter) of forceps minor; L Hipp = left hippocampus.

SU prediction was driven by a qualitatively different feature profile, with overlapping and distinct factors predicting PLEs compared with SU (Fig. 2). SU was most strongly predicted by elevated prenatal tobacco and alcohol exposure, family history of alcohol use disorder, religious prohibitions against alcohol and drug use, fun seeking, sensation seeking, drive, BIS, externalizing behaviors, and higher fluid and crystallized cognition. Lower income and parental education additionally predicted adolescent SU. Among neuroimaging features, higher auditory network to left-putamen rsFC and IFG activation during correct-go response execution (SST: correct-go vs. fixation) showed protective effects, while higher vmPFC activation to reward cues was associated with increased SU risk (Fig. 2; Supplementary Figure 5).

Strikingly, several features showed opposite directional effects across outcomes: higher fluid and crystallized cognition, DMN within-network rsFC, FPN-right nucleus accumbens rsFC, and sensation seeking, among the strongest predictors of SU, were in fact protective against PLEs, whereas IFG pars triangularis transverse diffusivity, among the strongest predictors of PLEs, was protective against SU (Fig. 2B). Baseline PLEs were also a strong predictor of SU risk, providing direct feature-level evidence for shared early vulnerability. Sex was associated with both outcomes such that females were at higher risk, and prenatal exposures, family history of drug and alcohol use, and religious preferences dominated SU prediction in a way that had no parallel in the PLE profile, suggesting that prenatal environment specifically shapes SU trajectories.

Directional SHAP feature importance profiles for each comparison are presented in Supplementary Figures 6 and 7.

### 3. A comorbidity risk signature predicts co-occurring psychosis spectrum symptoms and substance use

A distinct risk profile emerged for co-occurring outcomes, with partial overlap with the PLE and SU models. Predictors shared across all three outcomes included higher baseline PLEs, sex (females at increased risk), elevated externalizing behaviors, lower household income, higher positive urgency, higher fun seeking, higher ACEs, and higher family history of depression. Shared neuroimaging predictors included lower forceps minor TD and higher fusiform activation to negative faces on the eNBack task (Figs 3A-B).

**Figure 3.**
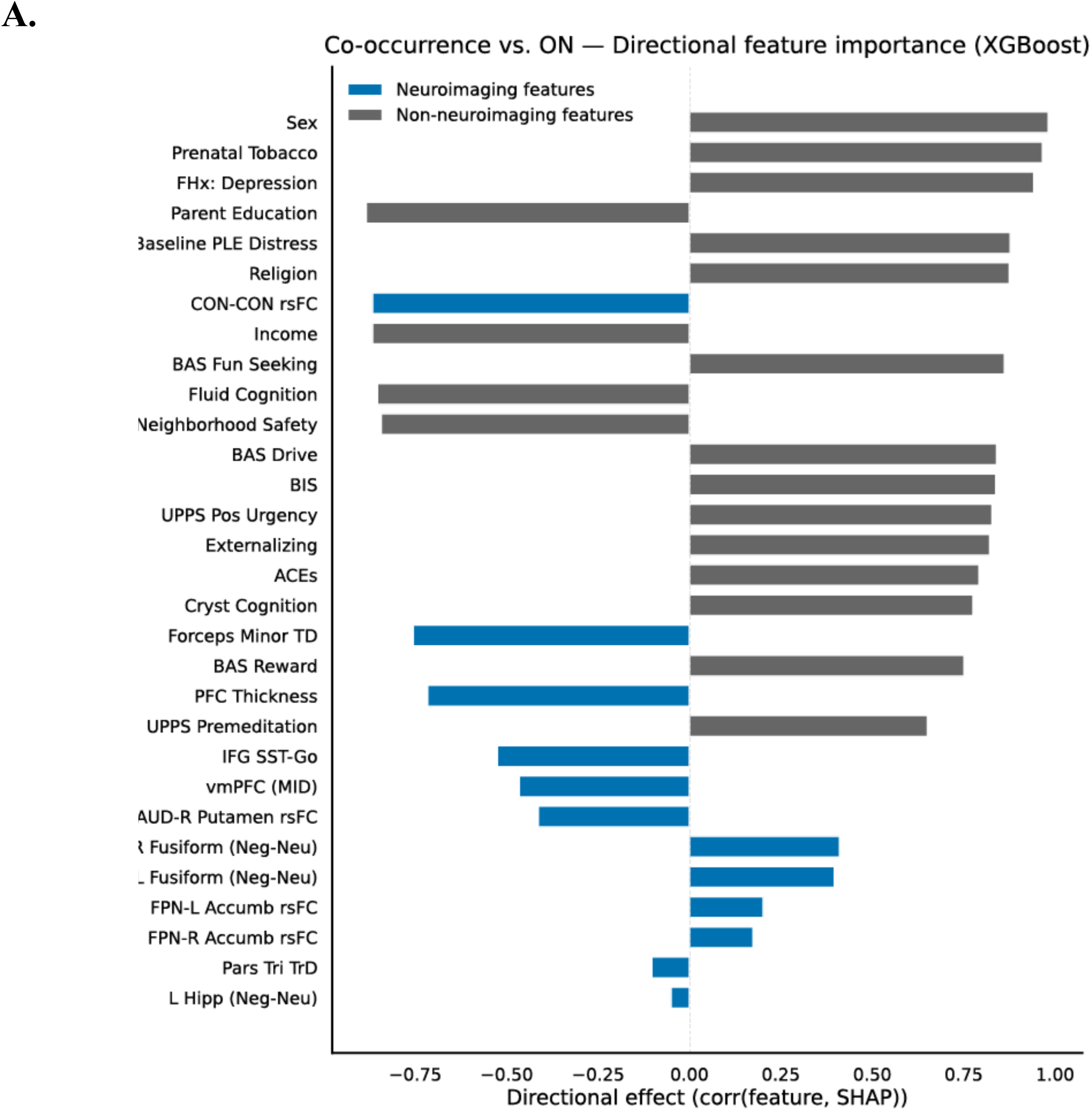
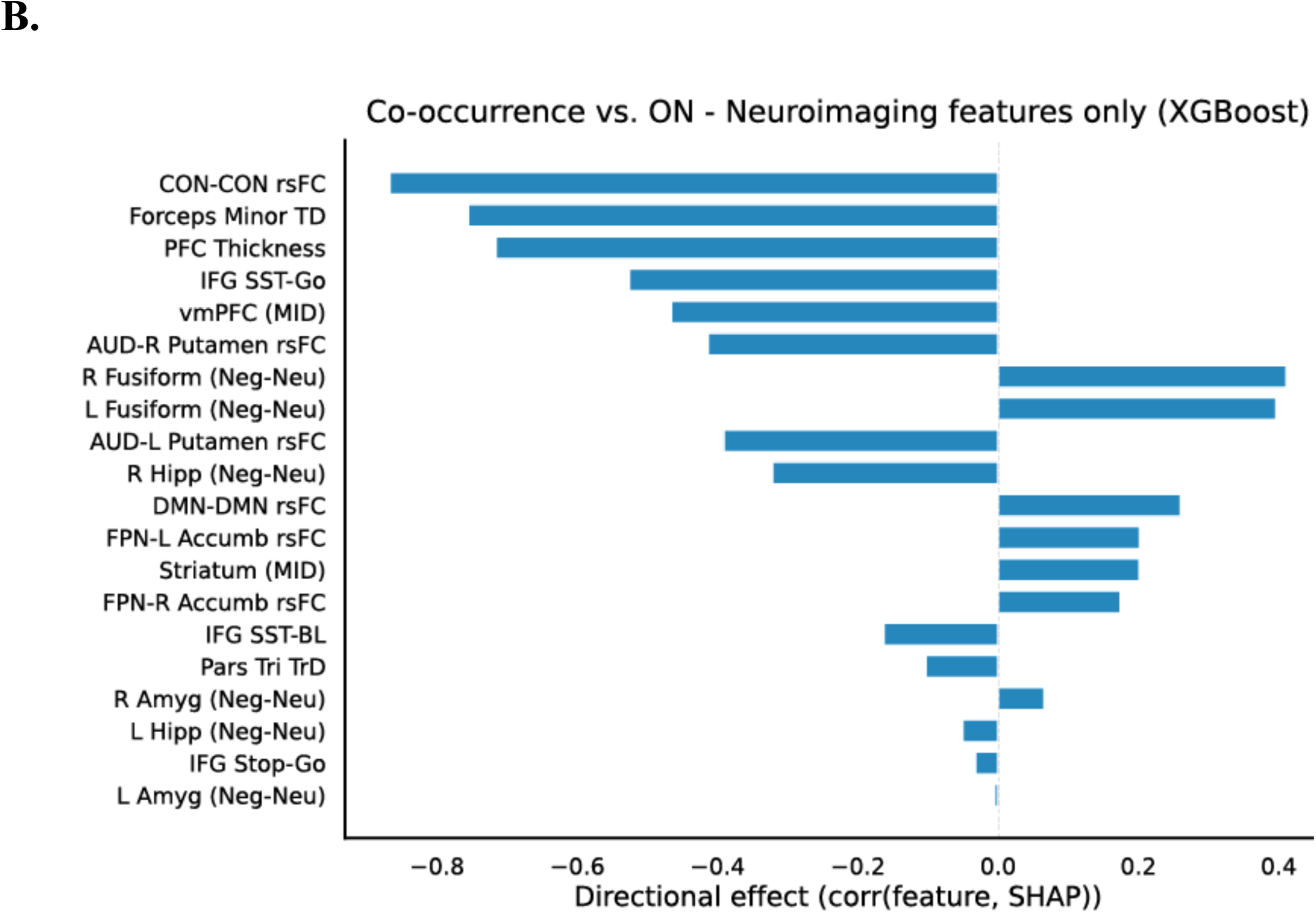
XGBoost directional feature importance for the Co-occurrence versus Outcome-Negative (ON) comparison. A). Directional feature importance rankings for top 30 most predictive features, derived from the signed correlation between each feature and its SHAP value on held-out test data, indicating whether higher feature values increase (positive) or decrease (negative) predicted probability of co-occurrence. Bars are color-coded by feature domain (blue = neuroimaging; gray = non-neuroimaging). B). Directional feature importance for neuroimaging features only, highlighting the relative contribution and direction of neuroimaging predictors. Together, panels A and B characterize the distinct comorbidity risk signature identified by the model. Directional labels (Higher / Lower) reflect the sign of the mean SHAP-feature correlation. Sex at birth was coded as binary; positive SHAP values indicate higher risk associated with female sex. **Abbreviations:** FHx = family history; CON = cingulo-opercular network; rsFC = resting-state functional connectivity; BAS = behavioral activation system; BIS = Behavioral Inhibition System; UPPS = UPPS Impulsive Behavior Scale; ACEs = adverse childhood experiences; Cryst cognition = crystallized cognition; Forceps Minor TD = restricted total diffusion (white matter) of forceps minor; PFC thickness = prefrontal cortical thickness; IFG = inferior frontal gyrus; IFG SST-Go = IFG activation during response inhibition (SST: inhibition vs. baseline); IFG Stop-go: IFG activation during response inhibition (SST: inhibition vs. go); IFG SST-BL = IFG activation during correct-go response execution (SST: correct-go vs. fixation); vmPFC (MID) = ventromedial prefrontal cortex activation during the Monetary Incentive Delay Task; AUD = Auditory Network; R fusiform = right fusiform gyrus; Neg-Neu = brain activation to negative faces (versus neutral faces) from the Emotional nBack Task; FPN = Frontoparietal Network; Accumb = nucleus accumbens; Pars Tri TrD = Transverse diffusivity (grey matter) of the IFG pars triangularis; L Hipp = left hippocampal activation to negative faces during the eNBack task; DMN = Default Mode Network; Striatum (MID) = striatal activation during the reward anticipation phase of the Monetary Incentive Delay Task; Amyg = Amygdala; MID = Monetary Incentive Delay Task.

Overlap specific to the co-occurrence versus PLE model included higher reward responsiveness (BAS), higher BIS, higher internalizing, and higher IFG activation during response inhibition (SST: inhibition vs. baseline). Overlap with the SU model included higher externalizing behaviors, prenatal tobacco exposure, and religious factors. Like in the substance use model, co-occurrence was predicted by higher bilateral frontoparietal network to nucleus accumbens rsFC.

Several features showed effects specific to co-occurrence or diverged from single-outcome models. Lower CON within-network rsFC was the strongest neuroimaging predictor of co-occurrence, but did not emerge as a predictor in PLE or SU models. Similarly, lower prefrontal cortical thickness and auditory network to right putamen rsFC, neuroimaging predictor of co-occurrence, did not emerge as predictors in PLE or SU models. In contrast to higher vmPFC activation to rewards observed in the SU model, vmPFC hypoactivation was observed in the co-occurrence model. While higher DMN within-network rsFC predicted higher SU in the SU model, but lower risk for PLEs in the PLE model, it was not a meaningful predictor in the co-occurrence model.

Higher drive towards reward, higher premeditation, and lower neighborhood safety were also uniquely predictive of co-occurrence. Sex showed the largest directional effect for co-occurrence exceeding its effect in either single-outcome model, with girls at greater risk compared to boys. Together, these findings indicate that co-occurring SU and psychosis-spectrum symptoms are not simply explained by additive risk from each domain, but instead reflect a partially distinct risk architecture with shared and unique neuroimaging and clinical contributors (Fig. 3A-B).

### 4. Co-occurrence is not reducible to either single-disorder profile

We next tested whether models trained to classify each group could directly discriminate between groups, addressing whether the strong co-occurrence classification performance observed in Results Section 2 might simply reflect the model leveraging the same psychosis-spectrum risk features used to predict PLEs. If co-occurrence were merely being classified based on shared PLE-related signal, we would expect models to fail at discriminating the co-occurring group from the PLE-only group directly. XGBoost models demonstrated above-chance classification across all three pairwise comparisons (Fig. 4A; co-occurrence versus SU-only: AUROC=0.765, SD = 0.030; co-occurrence versus PLE-only AUROC = 0.596, SD = 0.034; PLE-only versus SU-only AUROC=0.771, SD = 0.024). The ability to directly discriminate between all pairwise group combinations indicates that the co-occurring group carries a risk profile that is not reducible to psychosis-spectrum risk alone, consistent with the distinct comorbidity signature identified in Section 4. Directional feature importance profiles for each pairwise comparison are shown in Figure 4B. ROC curves for all secondary pairwise comparisons are shown in Supplementary Figure 8.

**Figure 4.**
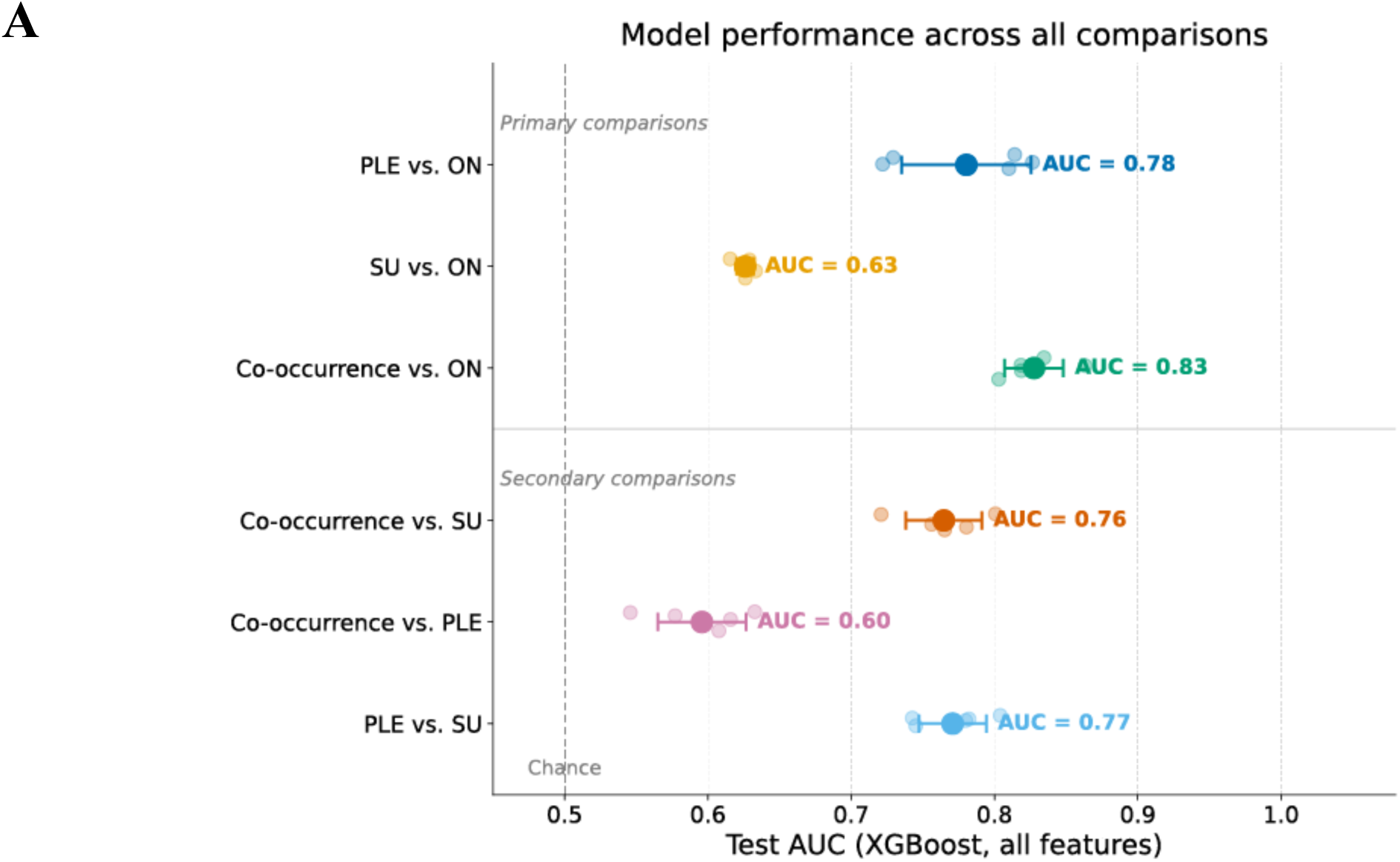
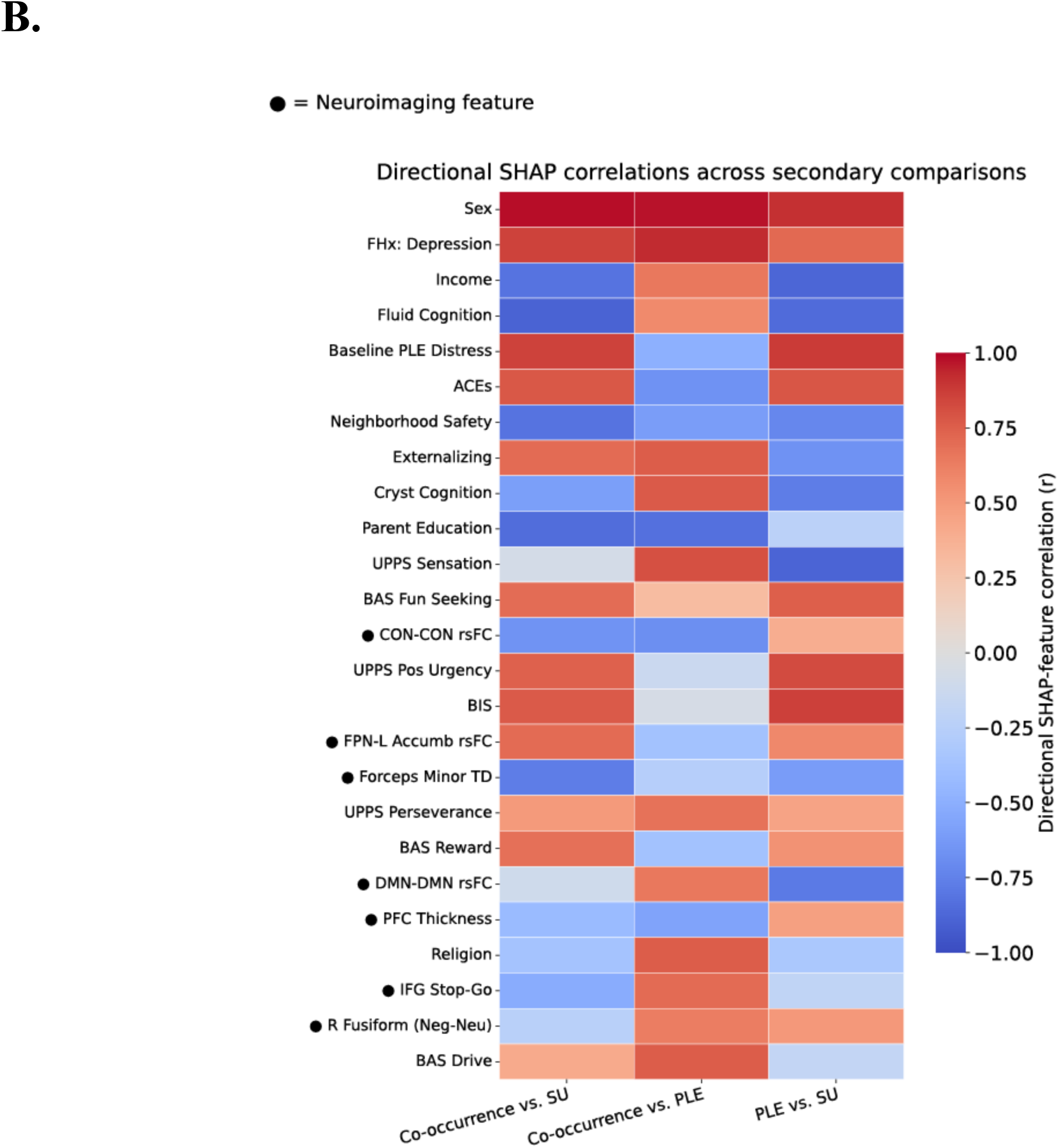
Secondary pairwise group discrimination analyses. A) XGBoost classification performance (mean AUROC ± SD across outer cross-validation splits) for all primary and secondary comparisons. Primary comparisons (top) show model performance for each single-outcome group versus outcome-negative (ON) youth. Secondary comparisons (bottom) show direct pairwise discrimination between outcome groups. Dashed vertical line indicates chance performance (AUROC = 0.50). B) Directional SHAP-feature correlations for the three pairwise secondary comparisons, showing the signed association between each feature and its SHAP contribution on held-out test data. Positive values indicate features associated with increased predicted probability of the first-named group; negative values indicate features associated with increased probability of the second-named group. Features are ranked by mean absolute directional effect averaged across all three pairwise comparisons. Sex at birth was coded as binary; positive SHAP values indicate higher risk associated with female sex. **Abbreviations:** FHx = Family history; ACEs = Adverse Childhood Experiences; Cryst cognition = crystallized cognition; UPPS = UPPS Impulsive Behavior Scale; BAS = behavioral activation system; CON = cingulo-opercular network; rsFC = resting-state functional connectivity; BIS = Behavioral Inhibition System; FPN = Frontoparietal Network; Accumb = nucleus accumbens; Forceps Minor TD = restricted total diffusion (white matter) of forceps minor; DMN = Default Mode Network; PFC thickness = prefrontal cortical thickness; IFG = inferior frontal gyrus; SST = Stop Signal Task; IFG Stop-go: IFG activation during response inhibition (SST: inhibition vs. go); R fusiform = right fusiform gyrus; Neg-Neu = activation to negative faces (versus neutral faces) from the Emotional nBack Task.

**Figure 5.**
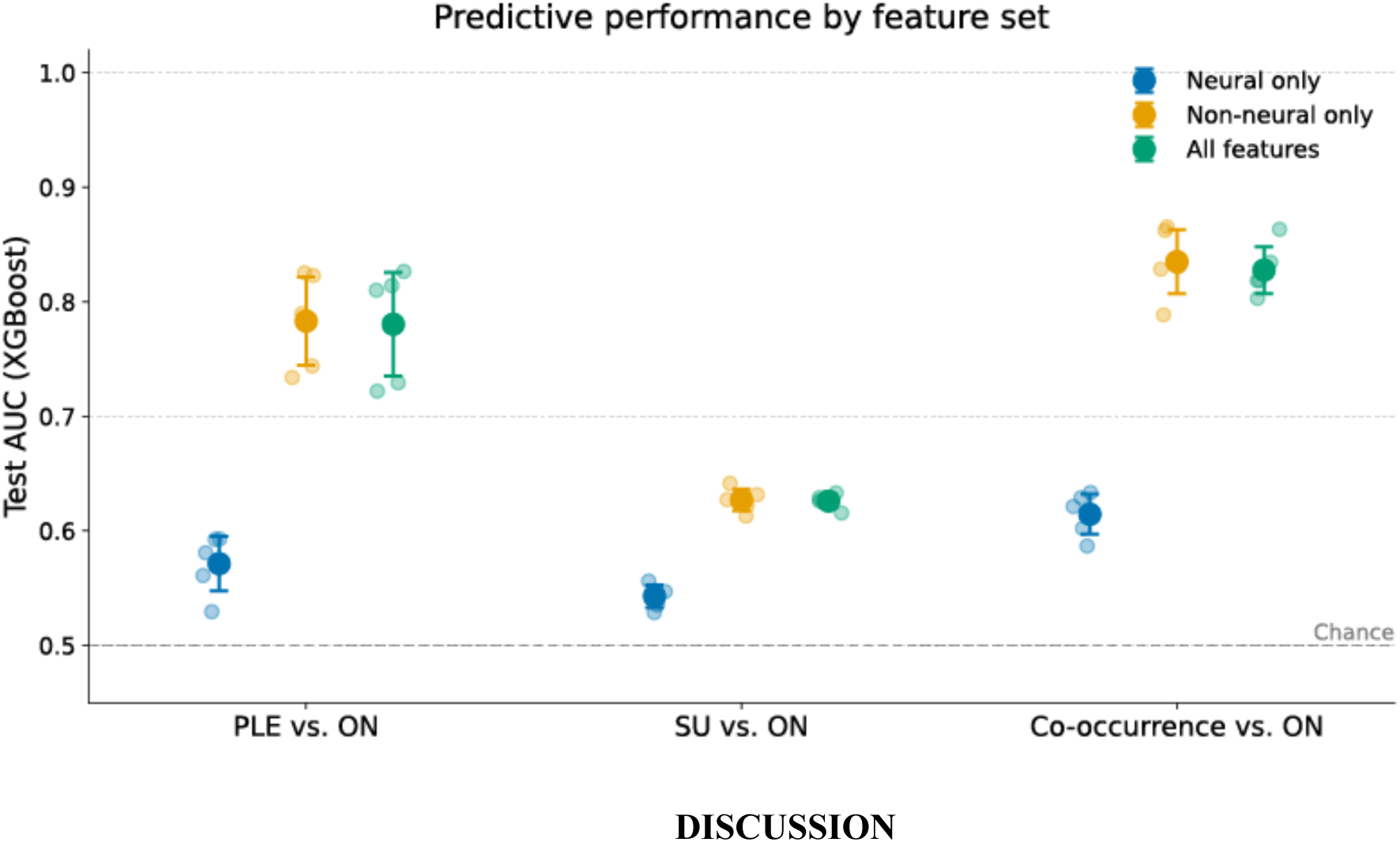
Neuroimaging feature contributions to outcome prediction. XGBoost classification performance (mean AUROC ± SD across outer cross-validation splits) for each primary outcome broken down by feature set: neuroimaging features only (neuroimaging), non-neuroimaging features only (demographic and clinical), and all features combined. Dashed horizontal line indicates chance performance (AUROC = 0.50).

### 5. Neuroimaging data alone accurately predict outcomes independently of demographic data

Finally, we examined whether neuroimaging features alone could predict outcomes independently of demographic and clinical information. Neuroimaging-only models demonstrated above-chance classification for SU and co-occurrence outcomes, with a trend-level effect for PLEs (Supplementary Figure 9; PLE vs. ON: AUROC = 0.575, SD = 0.027, permutation *p* = 0.062; SU vs. ON: AUROC = 0.543, SD = 0.011, permutation *p* = 0.003; Co-occurrence vs. ON: AUROC = 0.615, SD = 0.018, permutation *p* = 0.004; 1000 permutations each), indicating that baseline neuroimaging features carry an independent predictive signal beyond clinical and demographic factors. Among neuroimaging features, forceps minor total diffusion (TD) showed the greatest mean absolute SHAP contributions for both PLE and co-occurrence models. Next most informative were right fusiform activation to negative versus neutral faces for PLE, left auditory network-putamen rsFC and IFG pars triangularis transverse (radial) diffusivity for SU, and CON within-network rsFC and right auditory network–putamen rsFC for co-occurrence.

## DISCUSSION

Our study used multivariate machine learning models to predict adolescent substance use (SU), psychotic-like experiences (PLEs), and their co-occurrence from childhood demographic, clinical, and neuroimaging risk factors. Several novel findings emerged. First, childhood features prospectively predicted adolescent outcomes across all three classifications, indicating that vulnerability to SU and psychosis-spectrum symptoms is detectable years before symptom onset. Second, although some predictors were shared across outcomes, including two diffusion imaging features (IFG pars triangularis transverse diffusivity and forceps minor TD), each outcome showed distinct combinations of neuroimaging and environmental predictors, differing in both importance and directionality. These findings support partially dissociable developmental pathways for SU, PLEs, and their co-occurrence, rather than a single, shared liability. Third, co-occurrence was characterized by a distinct risk profile involving altered prefrontal structure and function, disrupted cortico-striatal connectivity and CON within-network rsFC, as well as reward system dysfunction. Finally, although clinical and demographic factors were the strongest predictors of outcomes, neuroimaging features provided independent predictive value, supporting their utility for biologically informed early risk identification. These findings advance an empirical foundation for early, biologically-informed identification of youth at risk for psychosis-spectrum and SU trajectories.

Notably, predictive accuracy was high despite outcomes reflecting very early and often low-level SU endorsement during adolescence. This suggests that meaningful neurobiological and environmental risk signatures are detectable well before the emergence of more severe or persistent psychopathology. Early SU risk and experimentation predict later SU disorders,^25,26^ and youth PLEs are associated with increased risk for adult psychotic disorders,^23,24^ highlighting the clinical importance of identifying vulnerability during early adolescence. Interestingly, prediction of SU alone was more modest, consistent with prior ABCD findings.^38^ Adolescent SU patterns are heterogeneous,^39^ and may reflect normative experimentation for some youth, whereas PLEs may reflect more enduring neurobiological vulnerability.^40,41^ The strong prediction of co-occurrence, while remaining distinguishable from PLE prediction, suggests that co-occurrence similarly reflects a relatively stable and biologically grounded phenotype.

Whereas prior studies in youth have modeled psychosis symptoms and SU separately,^42,43^ this represents the first study, to our knowledge, to identify premorbid predictors of co-occurrence in youth, and the first study in youth or adults to identify prospective neuroimaging predictors of co-occurrence. Altered prefrontal structure and function (i.e., lower prefrontal cortical thickness, elevated right IFG pars triangularis transverse diffusivity, lower forceps minor transverse diffusivity), and lower CON rsFC were among the strongest neuroimaging predictors of co-occurrence. Neuroimaging-only models similarly identified these prefrontal abnormalities, alongside additional task-based frontal alterations (inferior frontal gyrus activation during SST, vmPFC hypoactivation during the MID), as the strongest predictors of co-occurrence. This extends previous work linking white matter microstructure of the forceps minor and right IFG to prenatal cannabis exposure and PLEs,^44^ and work linking reduced PFC thickness to adolescent SU^42,45^ and PLEs^42^.

Importantly, several features appeared specific to co-occurrence. Reduced CON rsFC, lower prefrontal cortical thickness and auditory network-putamen connectivity, predicted co-occurrence but not PLE or SU individually, suggesting a distinct neurodevelopmental signature. Given the role of the CON in cognitive control.^46^, and evidence linking cortical thinning to psychosis conversion^47^ and addiction^48^, these convergent deficits may contribute to the particularly strong predictive performance observed for co-occurrence. Lower IFG activation during response inhibition predicting co-occurrence further implicated impaired prefrontal control mechanisms, consistent with prior work in youth at risk for psychosis.^49^

Co-occurrence was also characterized by dysregulated reward and motivational systems. Elevated trait-level impulsivity and externalizing symptoms were accompanied by vmPFC hypoactivation during reward valuation. Trait-level impulsivity predictors included greater reward sensitivity, positive and negative urgency, and reduced premeditation. Together with externalizing symptoms, these findings support a profile of heightened behavioral disinhibition. VmPFC hypoactivation during reward processing aligns with prior evidence of blunted reward-system engagement in youth PLEs, adult psychosis,^21,50^ and SU disorders.^51^ This pattern may reflect dissociations between self-reported motivational traits and underlying neural reward responses..^21,52,53^

Demographic and clinical predictors of co-occurrence largely converged with established risk factors for SU and PLEs. Lower household income, lower parental education, and higher ACEs predicted co-occurrence, consistent with extensive prior work linking socioeconomic adversity to both outcomes.^54–58^ Additional predictors included prenatal substance exposure, family history of alcohol use, and religion. We replicated previous findings showing that religious factors are protective against substance use.^38^

Cognitive predictors differentiated outcomes. Lower fluid cognition predicted greater risk, consistent with psychosis-spectrum vulnerability ^59,60^ whereas higher crystallized cognition predicted increased SU risk, paralleling prior work linking higher cognition to substance initiation in youth at clinical high risk for psychosis.^61^ Both contributed meaningfully to the prediction of co-occurrence, suggesting convergence of PLE-related and SU-related cognitive pathways.

Unexpectedly, female sex strongly predicted co-occurrence, with effects substantially exceeding those observed for SU or PLEs alone. Although adult schizophrenia^62^ and SU disorders,^63^ are generally more common in males, recent epidemiological data indicate rising SU rates among adolescent girls.^64^ he present findings may reflect heightened female vulnerability during adolescence related to pubertal timing,^65^ sex-dependent gene-environment interactions,^66^ and/or the narrowing sex differences in adolescent SU.^67–69^ Future studies should examine whether affective and stress-sensitive pathways contribute more strongly to co-occurrence risk in girls, whereas externalizing pathways in boys more often manifest as isolated SU trajectories. In previous machine learning studies using ABCD data, sex is often included as a covariate rather than evaluated as a directional predictor, which may obscure its predictive utility. ^70^

Several predictors overlapped across SU and PLE models, supporting transdiagnostic liability. Prefrontal dysfunction, including altered IFG activation during inhibitory control, predicted both outcomes, consistent with prior evidence linking impaired inhibitory control to psychosis risk and SU vulnerability.^71,72,73^ Baseline PLE severity was the strongest predictor not only of later PLEs, but also of SU and co-occurrence, suggesting that subthreshold psychotic experiences index broader developmental vulnerability rather than merely reflecting within-construct continuity.

Despite these shared predictors,important dissociations emerged. DMN rsFC showed opposing directional effects: lower DMN connectivity predicted PLEs, whereas higher DMN connectivity predicted SU and co-occurrence. The shared direction of DMN effects across SU and co-occurrence is consistent with prior work suggesting DMN dysfunction across both psychosis-spectrum and addiction phenotypes.^74,75^ Reduced DMN connectivity replicates prior ABCD findings in PLE prediction,^42^ while increased DMN connectivity in SU may reflect developmental patterns preceding disordered substance use.^74^

Reward-related activation also differed across outcomes. Frontal reward abnormalities appeared transdiagnostic, whereas subcortical reward hypoactivation was more prominent in SU consistent with substance-specific alterations in reward processing and reward-learning deficits.^76–79^ This frontal-subcortical reward dissociation also aligns with developmental models in which subcortical reward systems mature earlier than prefrontal control circuits.^77,80,81^

Methodologically, this study leverages gradient-boosted decision trees (XGBoost) with nested cross-validation, permutation testing, and SHAP-based interpretation to compare predictors across single-outcome and comorbid models. This framework enabled identification of both shared and outcome-specific predictors while preserving directional interpretability. While prior ABCD studies have used highly dimensional feature spaces,^38,42,82,83^ the current analyses achieved comparable performance using a relatively small curated set of theoretically-motivated features, supporting the value of developmentally grounded feature selection. Future work should extend this framework by modeling longitudinal trajectories of symptom emergence, evaluating generalizability in independent and clinically-enriched samples, and integrating mechanistic experiments to test the potentially causal contributions of certain risk factors that we have identified here to PLE, SU, and their co-occurrence.

Our analyses support the utility of extreme gradient boosting (XGBoost) for analyzing large, multimodal human neuroimaging datasets, with XGBoost achieving the highest classification performance in the majority of our analyses. XGBoost held a particularly pronounced advantage in neuroimaging-only feature sets, such that it achieved highest performance in 5 of 6 neuroimaging-only models. This pattern supports previous reports of gradient-boosted decision trees as especially well-suited to neuroimaging prediction, where signal often resides in nonlinear interactions that additive models such as elastic net and ridge regression fail to capture.^8^ Crucially, this preserves directional feature attribution required for inference, unlike AI algorithms such as convolutional neural networks.^84^

### Limitations

Neuroimaging measures, particularly task-related fMRI, are constrained by reliability limitations that may reduce predictive precision.^85,86^ Although rigorous quality control and preprocessing procedures were employed, measurement noise remains possible. Further, while previous work supports that substance experimentation in youth may reflect early vulnerability to later onset SU disorder,^25,26^ SU severity, frequency, and duration were relatively low in this cohort, and some early experimentation may reflect normative behavior rather than emerging disorder. This study was also underpowered to evaluate substance-specific effects (i.e., cannabis vs. alcohol vs. tobacco; Supplementary Table 7). Additionally, smaller co-occurrence and PLE group sizes may have reduced power despite balanced cross-validation procedures. Finally, causal inferences cannot be drawn from these associations.

## CONCLUSION

Collectively, our results demonstrate that childhood neuroimaging, behavioral, and environmental features can prospectively predict adolescent psychotic-like experiences, substance use, and their co-occurrence. Results underscore the importance of explicitly modeling comorbidity, as co-occurring trajectories may involve partially distinct neurodevelopmental mechanisms that are not apparent when SU and PLEs are studied separately.

## METHODS

This study was preregistered on the Open Science Framework prior to analysis (https://osf.io/y2n7j/files/fzyje). Although the ABCD dataset had already been collected, the preregistration specified study aims, outcome definitions, feature domains, preprocessing procedures, and modeling approaches. The present manuscript reports results from Aim 1 only, focusing on the prediction of adolescent substance use, psychosis spectrum outcomes, and their co-occurrence from baseline childhood risk factors. Preregistration deviations are reported in Supplementary methods, section 1.

### Section 1: Participants

Data were drawn from the Adolescent Brain and Cognitive Development (ABCD) Study, a longitudinal, multi-site investigation of neurodevelopment in 11,880 children initially recruited at ages 9–10 years across 21 U.S. research sites. The ABCD Study includes repeated assessments of demographic, clinical, behavioral, and neuroimaging measures collected at regular intervals throughout adolescence.

Analyses used data from ABCD Data Release 6.0. Baseline (Year 0) demographic, clinical, and neuroimaging features were used to predict outcome group status at follow-up study timepoints through Year 6 (mean age 16.1 years old at the Year 6 follow-up; SD = 0.66; n = 5,056 with data). See Supplementary Table 1 and Supplementary Methods, section 2 for further details. Baseline neuroimaging data were screened using ABCD-recommended quality control procedures. Data that passed functional neuroimaging quality control recommendations were included (https://docs.abcdstudy.org/latest/documentation/imaging/#quality-control-and-recommended-image-inclusion-criteria). Quality control metrics were implemented by domain (i.e., structural MRI, diffusion, and separately by task for fMRI) in line with the aforementioned recommendations. To further minimize potential sources of imaging-related variance, mean framewise displacement, outlier volumes, scanner type, and MRI serial number were regressed out of the data (see Section 5 Modeling methods for further details).

### Cohort descriptive statistics

After applying our filtering criteria (see Supplementary Figure 1 for sample construction flow chart), the final analytic sample comprised 10,134 participants: 281 PLE-only, 3,703 SU-only, 326 co-occurrence, and 5,824 outcome-negative (ON). Across the full sample, participants were on average 9.96 years old at baseline (SD = 0.62), 14.2 years old at the Year 4 follow-up (SD = 0.71; n = 9,735 with data), and 16.1 years old at the Year 6 follow-up (SD = 0.66; n = 5,056 with data). The sample was 52.5% male (n = 5,322) and 47.5% female (n = 4,812) (Supplementary Table 1).

### Section 2: Outcome group definitions

Four mutually exclusive outcome groups were derived from adolescent follow-up data collected across post-baseline assessment waves (Supplementary methods, section 2). Psychotic-like experiences (PLE) were assessed using the Prodromal Questionnaire – Brief Child Version (PQ-BC), with distress scores calculated as the sum of 21 items weighted by endorsed level of distress (range: 0–126). Consistent with Karcher et al. (2024)^42^, a wave-specific threshold was computed at each assessment wave as 1.96 standard deviations above that wave’s mean distress score. Youth were classified as PLE-positive if they met or exceeded this threshold at two or more annual post-baseline assessment waves (Years 1 through 6), reflecting a persistence criterion designed to identify youth with sustained, clinically meaningful psychosis-like experiences across development rather than isolated or transient elevations.

Substance use (SU). Substance use was assessed using a research-assistance-administered substance use Timeline Follow-Back (TLFB) interview and Initial Substance Intake Patterns (iSIP) screener administered at each study wave; TLFB and iSIP administration methods have been previously described.^87,88^ Endorsement of alcohol, nicotine, or cannabis use was evaluated, with substance use defined as “full use” (≥1 standard alcoholic drink or more than a puff/taste of cannabis or nicotine, or other occasion of substance use).^89,90^ We also analyzed hair, urine, salivary, and breathalyzer toxicology measures (Supplementary methods, section 3; Supplementary Tables 5 and 6; Supplementary Figure 8). Youth who screened positive for any substance were included in the SU group regardless of self-report. Among participants who reported no substance use, 14.15% tested positive on toxicology at Year 4 and 20.67% at Year 6, a pattern consistent with the literature on increased underreporting of substance use as adolescents age.^90^ The final SU outcome was defined as self-reported endorsement of full use on the TLFB interview or the iSIP screener, or a positive toxicology test across any modality at Year 4 or Year 6. This operationalization was adopted as the primary definition based on measurement validity and alignment with the preregistered analytic intent; four alternative operationalizations were considered but not pursued, as discussed in Supplementary Methods and Results, section 2, *Substance use outcome definition*.

Co-occurrence group. Co-occurrence was defined as meeting criteria for both PLE and SU outcomes. This group was mutually exclusive from the PLE and SU groups; that is, a participant meeting criteria for both the PLE and SU definitions would be included in the co-occurrence group, but not the PLE-only or SU-only groups.

Outcome-negative. Participants who did not meet criteria for any of the other outcome groups are referred to as ‘Outcome-negative’ in descriptive and comparative analyses.

A total of 1,746 participants were excluded because they had no available substance use data (no self-report substance use data and no toxicology) at either the Y4 or the Y6 follow-up wave. The final analytic sample consisted of 10,134 participants (Co-occurrence: n = 326; PLE-only: n = 281; SU-only: n = 3,703; ON: n = 5,824). Due to the extreme class imbalance between positive outcome groups and the ON group, the ON group was downsampled to a 5:1 ratio for the PLE-only and co-occurrence models before model training (Supplementary methods and results, section 6; Supplementary Table 8). The SU-only model used the full dataset, given the more favorable natural class ratio (approximately 1.6:1), with class imbalance addressed via the ‘scale_pos_weight’ parameter in extreme gradient boosting (XGBoost). XGBoost, by design, implements built-in handling of imbalance through its ‘scale_pos_weight’ function. This may be statistically preferable to pre-matching, which can inadvertently discard informative variation.^91^ ‘scale_pos_weight’ increases the penalty for misclassifying minority-class examples so that the model learns balanced decision rules without altering the underlying sample. Our XGBoost analysis workflow, therefore, does not utilize matching; instead, we applied a stratified 80/20 train-test split, and adjusted imbalance exclusively within the training data.

### Section 3: Demographic and clinical input features

We first tested whether a relatively small set of theoretically-informed baseline demographic, clinical, and neuroimaging features predicted adolescent psychosis spectrum symptoms and SU endorsement. Demographic and clinical features included sex, household income, parental education, religious factors, neighborhood-level factors, childhood adversity, family conflict, prenatal substance exposures, family history of psychopathology, baseline psychopathology symptoms, impulsivity facets, and behavioral activation and inhibition facets (see Methods, section 4 for a complete list of features). Neuroimaging features included task-related functional magnetic resonance imaging (fMRI), including activation of the inferior frontal gyrus (IFG) from the Stop Signal Task (SST), activation in reward-related regions (e.g., striatum) from the Monetary Incentive Delay (MID) Task, amygdala, hippocampus, and fusiform gyrus activation during the Emotional N-back Task (eNBack), structural magnetic resonance imaging (MRI) measures including prefrontal cortical thickness, resting-state functional connectivity (rsFC) measures including default mode network (DMN) within-network rsFC, cingulo-opercular network (CON) within-network rsFC, bilateral frontoparietal network (FPN)-to-accumbens connectivity, and bilateral auditory network-to-putamen connectivity. This theoretically-grounded set of features was selected for developmental and clinical relevance to substance use and psychosis outcomes (Methods, sections 4 and 5). A total of 53 predictors (across demographic, clinical, and neuroimaging predictors) collected at baseline were considered, a notably smaller number than previous ABCD prediction studies.^38,42,82,83^ For a complete inventory of all baseline features used in the prediction models, see Supplementary Methods, section 4. Features were selected *a priori* based on ABCD, developmental psychopathology, and substance use literatures.

Baseline demographic features (eight in total) included: participant sex, household income, parental education level, neighborhood safety^38^, school detention or suspension history^38^, religious affiliation^38^, religious prohibitions against alcohol use^38^, and religious prohibitions against drug use^38^. Clinical features (25 in total) included: adverse childhood experiences (ACEs) derived from the Kiddie Schedule for Affective Disorders and Schizophrenia (KSADS)^92^ as in Karcher et al, 2024^42^, family conflict as in Ho et al, 2022^93^, internalizing symptoms^94^, externalizing symptoms^95^, prenatal cannabis, alcohol, and tobacco exposure status^96^, parental history of mental illness (alcohol use problems, drug use problems, depression, mania, and psychotic symptoms)^42,93^, trait-level impulsivity measured using the UPPS Impulsive Behavior Scale (negative urgency, positive urgency, perseverance, premeditation, and sensation seeking)^97^, behavioral inhibition and activation system sensitivity measured using the BIS/BAS scales (BIS, BAS drive, BAS fun seeking, BAS reward responsiveness)^98^, baseline PLEs, baseline substance use, and cognitive performance measured using the National Institutes of Health (NIH) Toolbox fluid and crystallized cognition composites.^99^

Demographic and clinical features were examined both independently and in combination with neuroimaging features (see Section 4) to evaluate their relative and additive contributions to predictive performance (Supplementary Table 2B); collinearity between sociodemographic features was modest (all pairwise *rs* ≤ .66 (Supplementary Figure 9).

### Section 4: Neuroimaging input features

#### Imaging Procedure: Processing

ABCD imaging data collection, acquisition, and analysis have been previously described.^100–102^ The ABCD Data Analysis, Informatics, and Research Center (DAIRC) performs centralized processing of neuroimaging data from the ABCD study using the multi-modal processing stream.^103^ We included pretabulated derived variables from structural MRI, resting-state fMRI, and task-related fMRI from three tasks: the Monetary Incentive Delay (MID), Emotional n-Back (eNBack), and Stop Signal (SST) tasks. Resting-state data comprised four 300-second runs per participant, and task-related fMRI comprised two runs per task (MID: 322.4 s; n-Back: 289.6 s; SST: 349.6 s). A complete inventory of all baseline features used in the prediction models, including domain, variable name, ABCD source variable(s), derivation method, and description is provided in Supplementary Table 4 (ABCD Data Release 6.0. Baseline, Year 0).

#### Resting-State Functional Connectivity Methods

After ABCD DAIRC processing, between-network connectivity was calculated by computing pairwise correlations between each region of interest within given network and each region of interest within another network, defined by the Gordon parcellation.^104^ These correlations were averaged and Fisher *z* transformed to generate a summary metric of between-network connectivity strength.

#### Structural Magnetic Resonance Imaging Methods

Participants completed T1- and T2-weighted structural scans (1 mm isotropic) on a 3T scanner (Siemens, General Electric, or Phillips) with a 32-channel head coil. Structural neuroimaging data were processed using FreeSurfer version 5.3.0 through ABCD’s standardized processing pipelines.^100^ Average prefrontal cortical (PFC) thickness was computed as the mean of the bilateral caudal middle frontal, rostral middle frontal, and superior frontal gyri.^42^

#### Task-related Activation Methods

We additionally utilized summary metrics of task-related activation available in the tabulated data, computed by the ABCD DAIRC using generalized linear models: specifically, we utilized normalized beta coefficients for a given contrast (see list of Baseline features, below, Supplementary Table 4, and doi: 10.1016/j.neuroimage.2019.116091).103

#### Baseline Neuroimaging Features

Baseline neuroimaging features (20 in total) spanned structural MRI, diffusion MRI, resting-state functional connectivity, and task-related fMRI measures, preregistered based on previous studies indicating predictors of relevant mental health outcomes in the ABCD study, as well as previously reported robust general predictors of psychopathology and/or substance use. Structural features included average PFC thickness^42^ and diffusion-derived measures of white and gray matter microstructure including restricted total diffusion (white matter) of forceps minor^44^ and transverse diffusivity (grey matter) of the IFG pars triangularis^44^ (see Fig.6, Baseline Features list, below, and Supplementary Table 4). Resting-state features included within-network connectivity of the default mode and cingulo-opercular networks,^42^ as well as frontostriatal and auditory network connectivity.^44,105^ Task-related activation measures were extracted from the SST^101,106^, MID^101,107^, and eNBack^101^ using *a priori* regions of interest implicated in inhibitory control, reward processing, and emotion regulation (see Fig.6, Baseline Features list, below).21,49,51,108

Neuroimaging features were analyzed both as a standalone predictor set and in combination with demographic and clinical features to assess modality-specific predictive utility.

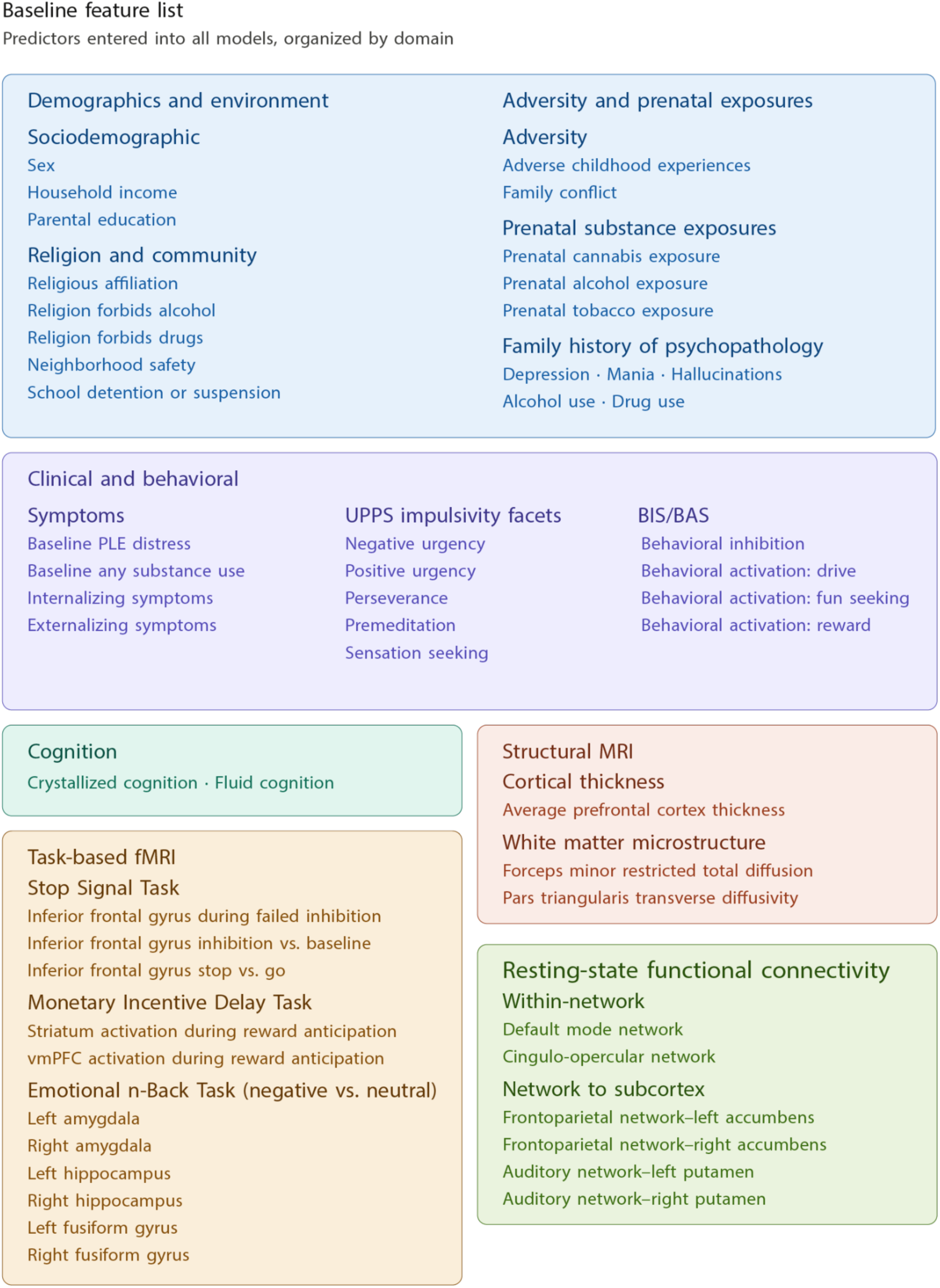

Baseline Features list. Complete list of baseline features. An inventory of all baseline features used in the prediction models, including domain, variable name, ABCD source variable(s), derivation method, and description is provided in Supplementary Table 4. **Abbreviations:** PLE = psychotic-like experiences; MRI = magnetic resonance imaging; fMRI = functional magnetic resonance imaging; vmPFC = ventromedial prefrontal cortex.

### Section 5: Modeling methods

Across our analyses, we tested six binary comparisons: our primary models compared each outcome group against the outcome-negative (ON) group (PLE-only vs. ON, SU-only vs. ON, and Co-occurrence vs. ON), and three binary comparisons as secondary analyses (Co-occurrence vs. SU-only, Co-occurrence vs. PLE-only, and PLE-only vs. SU-only). To quantify the relative contribution of neuroimaging versus non-neuroimaging predictors, each comparison was repeated across three feature sets: all features combined, neuroimaging features only, and non-neuroimaging features only, yielding 18 total model configurations. Within each configuration, we evaluated five supervised classification algorithms (XGBoost, random forest, ridge, elastic net, and SVC), consistent with the preregistered analysis plan. Permutation tests (1,000 label permutations) were applied to the all-features XGBoost model for each of the six comparisons, and SHAP values were computed across all 18 XGBoost configurations to support feature-importance interpretation.

For all structural and functional MRI features, we first regressed out the effects of site, scanner type, and MRI serial number (after dummy coding each of these categorical variables) using ordinary least squares linear regression; for all functional MRI features, mean framewise displacement was also regressed out.^93^ For all continuous features, missing values were imputed using the median value of the respective feature; for all categorical features, missing values were imputed using the modal value of the respective feature.^93^ Critically, data were imputed after splitting the data to prevent the risk of leakage between the training/validation and test sets. For XGBoost, missing values were not imputed because the algorithm natively models missingness during split optimization; imputation was therefore restricted to linear and distance-based models that require complete feature matrices.

Model performance was evaluated using stratified 5-fold cross-validation, which preserves class balance across folds and provides an unbiased estimate of out-of-sample performance (see Supplementary methods, section 5 for full details on preprocessing pipeline). All preprocessing steps (including scaling) were performed within each training fold to prevent information leakage, using Python scikit-learn pipelines. For ridge, elastic net, and SVC, hyperparameters were tuned using nested cross-validation, with an inner GridSearchCV loop for model selection and an outer loop of repeated stratified train/test splits (5 random seeds) for performance estimation (Supplementary methods, section 6). Random forest, ridge, elastic net, and SVC models were therefore all assessed using held-out test predictions, providing a rigorous and leakage-free estimate of model generalization. We additionally employed XGBoost classification. Given XGBoost’s substantially larger Bayesian-optimized hyperparameter space, we used repeated stratified train–test splits to obtain a stable distribution of held-out performance estimates; the comparison models, with smaller grid-searched hyperparameter spaces, were evaluated via standard nested cross-validation. Specifically, XGBoost was evaluated using repeated stratified train–test splits (5 iterations; 80% training, 20% test), with hyperparameters tuned via Bayesian optimization (75 iterations) using stratified 5-fold cross-validation within each training split. Permutation tests were conducted on a single stratified 80/20 train/test split with 1,000 label permutations. Best-performing hyperparameters for each comparison are reported in Supplementary Table 9.

Multiple supervised classification algorithms were evaluated, consistent with the preregistered analysis plan. These included penalized logistic regression models (ridge and elastic net regularization), SVC, random forest classifiers, and XGBoost. Model performance was compared across algorithms and feature sets. Performance is reported as the mean test AUROC ± standard deviation across splits (with range), with all metrics derived from held-out test sets in the outer loop, and SHAP values are interpreted from the highest-performing split. Receiver operating characteristic (ROC) curves are plotted from a best-performing model fit from a single outer split for visualization (Results section 1), whereas model performance in subsequent main text and tables is reported as the mean test area under the curve (AUROC ± standard deviation and range) across 5 independent outer train–test splits to provide a robust estimate of generalization performance. Given low base rates, class imbalance, and the dimensional overlap of early outcomes, we used parallel binary classifiers for each outcome rather than multiclass models to preserve sensitivity and interpretability (see Supplementary methods, section 6, “Rationale for Binary Outcome versus Outcome-Negative Classification Approach”).

Finally, we conducted a series of validation analyses to assess whether observed model performance could be attributed to confounding factors, data leakage, or spurious structure in the data; we also re-ran models without baseline PLEs as an input feature, and found that model performance was essentially unchanged (Supplementary methods and results, section 7).

### Section 6: Characterization of predictor variable associations

We used Shapley additive explanations (SHAP) to interpret variable importance in the model.109,110 SHAP values provide an interpretable measure of each feature’s contribution to the predicted outcome for each participant, expressed in the units of the model’s output. For our binary classification models, this is log-odds: for example, a SHAP value of −1 for family income means that participant’s income decreased their predicted log-odds of the outcome by 1, relative to the model’s baseline expectation.

## Supporting information

Supplementary Material Main

## Data Availability

The data used in this study were obtained from the Adolescent Brain Cognitive Development (ABCD) Study, a publicly available dataset managed by the National Institutes of Health (NIH). Researchers can request access to the ABCD Study data through the NIH Brain Development Cohorts (NBDC) Data Access Committee https://www.nbdc-datahub.org/data-access-process. Access is restricted to qualified researchers affiliated with an institution, and approval is contingent on compliance with the NBDC Data Use Certification, which is available at https://cdn.prod.website-files.com/672cec1c3c6a5c9ee60d1184/6852ddeaf0589dda767af337_General%20Individual%20v1.0.0.pdf. Data can be used for scientific research purposes only and cannot be used for commercial or nonresearch applications.

## Acknowledgements

This work is supported by the National Institute on Drug Abuse (Grant No. F31DA060068 [to CMA]).

Data used in this study were obtained from the Adolescent Brain Cognitive Development (ABCD) Study (https://abcdstudy.org). The ABCD Study is supported by NIH and additional federal partners under award numbers U01DA041048, U01DA050989, U01DA051016, U01DA041022, U01DA051018, U01DA051037, U01DA050987, U01DA041174, U01DA041106, U01DA041117, U01DA041028, U01DA041134, U01DA050988, U01DA051039, U01DA041156, U01DA041025, U01DA041120, U01DA051038, U01DA041148, U01DA041093, U01DA041089, U24DA041123, and U24DA041147. A full list of supporters is available at https://abcdstudy.org/federal-partners.html. A list of participating sites and a complete list of the study investigators is available at https://abcdstudy.org/consortium_members/. ABCD consortium investigators designed and implemented the study and/or provided data but did not necessarily participate in the analysis or writing of this report. The contents of this article are solely the responsibility of the authors and do not necessarily represent the official views of NIH or the ABCD consortium investigators.

A previous version of this article was published as a preprint on medRxiv.

## Code Availability

Relevant code is available at https://github.com/amircarolyn1/abcd-psychosis-su-prediction-pipeline.

## Author Contributions

Conceptualization including contribution to preregistration was done by all co-authors. Data processing was performed by CMA. Formal analysis was carried out by CMA. Code contributions were made by CMA, SEC, CW, and HRW. Code review was performed by CW. Manuscript preparation was performed by CMA. Review and editing was carried out by all co-authors. Supervision was provided by CEB.

