## Supplementary Material Main for "Predicting Substance Use and Psychotic-Like Experiences in Adolescents"

**Supplementary Materials**

### Table of Contents

**Supplementary Methods and Results**
1. Pre-Registration Deviations and Rationale
2. Outcome Group Construction
3. Toxicology Measures (Hair, Urine, Salivary, and Breathalyzer)
4. Baseline Input Features
5. Preprocessing Pipeline (Residualization, Standardization, and Missing Data)
6. Model Specification and Hyperparameters
7. Validation and Sensitivity Analyses
8. SHAP Value Computation and Interpretation

**Supplementary Tables**
Supplementary Table 1: Participant Demographics by Outcome Group
Supplementary Table 2: Per-Fold and Per-Seed AUROC Values
Supplementary Table 3: Negative control analysis using random labels
Supplementary Table 4: Full List of Baseline Input Features
Supplementary Table 5: Summary of substance use measures and variables
Supplementary Table 6: Self-Report vs Toxicology Concordance
Supplementary Table 7: Polysubstance use rates
Supplementary Table 8: Sample Sizes and Downsampling Parameters by Comparison
Supplementary Table 9: XGBoost and Comparison Model Hyperparameters

**Supplementary Figures**
Supplementary Figure 1: Sample construction flow chart
Supplementary Figure 2: Model Performance Stability Across Cross-Validation Folds and Random Seeds
Supplementary Figure 3: Precision-Recall Curves
Supplementary Figure 4: Calibration Plots Across Comparisons
Supplementary Figure 5: Directional feature importance for PLEs and SU
Supplementary Figure 6: SHAP Beeswarm Plots for Top Predictors by Comparison
Supplementary Figure 7: SHAP Mean Absolute Value Bar Plots by Comparison
Supplementary Figure 8: ROC Curves for Secondary Models
Supplementary Figure 9: Neuroimaging-only Permutation Tests
Supplementary Figure 10: Toxicology positivity by modality
Supplementary Figure 11: Feature Correlation Structure Within Sociodemographic Features

**Supplementary Methods 1:** Pre-Registration Deviations and Rationale

The pre-registration was filed with the project's registry following data collection but prior to analysis and is available at https://osf.io/y2n7j/files/fzyje. Below, we document each deviation from the pre-registered protocol with an accompanying rationale for each change.

Outcome Group Definitions

Pre-registered (SU): "Full use" of alcohol, nicotine, or cannabis defined as ≥ 1 standard alcoholic drink, > a puff/taste of cannabis or nicotine, or any other occasion of consumption/use, following Sullivan et al, 2022.

Implemented (SU): Self-reported "full-use" on the Timeline Followback (TLFB) and/or the Initial Substance Intake Patterns (iSIP) screener of any substance OR a positive toxicology screen at the Year 4 OR Year 6 follow-up wave. Toxicology positivity was determined from urine, hair, oral fluid, or saliva assays for any substance.

Rationale: Self-report alone substantially underestimates adolescent substance use, with our analytic sample showing a 14.1% false negative rate at Y4 and 20.7% at Y6 (toxicology-positive among self-report negatives; see Supplementary Table 6). Inclusion of toxicology screens substantially improves outcome ascertainment and reflects best practice in adolescent substance use research. Sensitivity analyses using a stricter "more than one day of use" definition yielded similar results (see Supplementary Methods 2).

Analytic Aims and Modeling Approaches

Stacked modeling was not implemented. The pre-registration proposed transmodal/stacked modeling combining single-channel modality predictions in a second-layer model. After preliminary work, this approach was deprioritized for the present manuscript because the analytic sample size, particularly for the rare PLE-only and Co-occurrence groups was insufficient to support both an inner cross-validation loop for first-layer models and a second-layer fitting procedure without substantial overfitting. Additionally, given the modest single-modality performance of neuroimaging features alone), the marginal gain from stacking was unlikely to justify the additional model complexity.

MVPA was not implemented. The pre-registration proposed ROI-based multivariate pattern analysis (MVPA) of reward-related regions (striatum, vmPFC) from the Monetary Incentive Delay Task. MVPA was deprioritized, following recent papers reporting low reliability for individual-level ABCD task fMRI data (Demidenko et al, 2024, *Imaging Neuroscience*, and Demidenko et al, 2024, *Dev. Cog. Neuro*). We therefore utilized univariate measures from the striatum and vmPFC to bolster reliability and stability.

Inference Criteria

Pre-registered: "Accuracy, Recall (true positive rate), AUROC, Weighted F1 scores, Precision."

Implemented: We reported area under the ROC curve (AUROC) as the primary performance metric, with sensitivity, specificity, positive predictive value, negative predictive value, and Brier score reported in supplementary materials. We additionally report permutation tests to establish the empirical null distribution and to compute *p*-values for primary comparisons; per-fold AUROC distributions and bootstrap confidence intervals were used for stability assessment (see Supplementary Table 2).

Rationale: AUROC is the most commonly reported summary metric for binary classification with class imbalance.

Feature Importance and Interpretation

Pre-registered: "Magnitude of each coefficient; SHAP values; Other explainability metrics may also be utilized for interpretability."

Implemented: SHAP (SHapley Additive exPlanations) values are reported as the primary feature importance metric. We computed SHAP values for each XGBoost model (TreeExplainer algorithm). Per-feature mean absolute SHAP values are reported, with directional SHAP values (signed by the Spearman correlation between feature values and SHAP contributions) used to indicate the direction of each feature's effect. XGBoost is a tree-based ensemble without comparable coefficient outputs and coefficient magnitudes are therefore not reported.

Cross-Validation Structure

Pre-registered: "Stratified cross-validation will be implemented in flat models to account for imbalanced group sizes."

Implemented: We utilized the following nested cross-validation methods:

- Outer loop: 5 outer train/test splits using different random seeds, each implementing an 80/20 stratified train/test split. Each outer split provides an independent estimate of held-out test AUROC, and we report the mean and standard deviation across the 5 splits are reported as the primary performance metric.
- Inner loop: Within each outer training set, we used 5-fold stratified cross-validation for hyperparameter optimization via Bayesian optimization (75 iterations per outer split), with random search as an automatic fallback if Bayesian optimization failed to converge.
- Per-split outputs: we report AUROC, sensitivity, specificity, positive predictive value, negative predictive value, and Brier score per outer split (Supplementary Table 2).

**Rationale:** A single train/test split, even with stratified cross-validation for hyperparameter tuning, yields a performance estimate that depends heavily on which participants happen to be assigned to the test set, particularly for the rare-positive comparisons (PLE-only and Co-occurrence). The 10-outer-split scheme provides a distribution of held-out performance estimates that better characterizes generalization variability and produces appropriately calibrated confidence intervals. The use of an inner 5-fold CV loop for hyperparameter selection ensures that hyperparameter choices are not contaminated by the held-out test data of the corresponding outer split. This nested structure is standard practice for unbiased performance estimation in machine learning research.

Addition of features: A study predicting substance use in adolescents from baseline risk factors was published after our preregistration (Green et al, 2026). We included additional feature predictors that we had not pre-registered, including: child's religious affiliation, religious prohibitions against alcohol use, religious prohibitions against drug use, UPPS, and BIS/BAS after the aforementioned study highlighted their relevance to substance use prediction.

Summary

Pre-registration deviations described above were either refinements that align with the pre-registered intent, methodologically driven choices intended to improve the validity or interpretability of results, or deferrals of secondary analyses due to methodological concerns. Overall, the primary aim, the data source, the predictor feature set, and the outcome structure all match the pre-registered protocol.

**Supplementary Methods 2:** Outcome Group Construction

Participants were classified into one of four mutually exclusive outcome groups based on their psychotic-like experience (PLE) and substance use (SU) trajectories across the follow-up waves of the ABCD Study. Group definitions are described below.

Psychotic-like experiences (PLEs). PLE status was determined using the Prodromal Questionnaire – Brief Child Version (PQ-BC) distress score. At each annual post-baseline assessment wave (years 1-6), participants were classified as meeting the PLE threshold if their distress score fell at or above 1.96 standard deviations above the wave-specific mean, corresponding to approximately the top 2.5% of the distribution at that wave. The wave-specific standardization was adopted to account for developmental changes in the distribution of PQ-BC scores across adolescence. A participant was included in this group if they met this criterion at two or more of the years 1-6 annual follow-up waves (meeting criteria for “persistent, distressing” PLEs, in line with Karcher et al, 2024).

Substance use (SU). SU status was determined using a combined self-report and toxicology approach (see Toxicology Measures). At each of the Y4 and Y6 follow-up waves, a participant was classified as having used substances if they endorsed any substance use on the Timeline Followback (TLFB) interview or iSIP screener, or screened positive on any hair, urine, salivary, or breathalyzer measure at that wave. A participant was classified as "full-use" for SU if they met this criterion at either Y4 or Y6. SU was not assessed at earlier years given the extremely low base rates of substance initiation at earlier time points.

The SU outcome was operationalized as “full use” of any substance (Sullivan et al, 2022) at the Y4 or Y6 follow-up wave, defined by self-reported use on the TLFB interview or iSIP and/or a positive toxicology screen at either wave. This definition was selected based on three considerations: 1) developmental framing: this study aimed to identify any adolescent substance exposure, consistent with the broader literature on adolescent SU initiation as a developmentally meaningful event (Hardee et al, 2018; Jackson et al, 2015; Magid et al, 2014; Behrendt et al, 2009); 2) measurement validity: combining self-report with drug toxicology reduces misclassification from documented adolescent underreporting (false negative rates of 14.1% at Y4 and 20.7% at Y6 in our sample; see Supplementary Table 6); and 3) alignment with the pre-registered intent. Our pre-registration specified "full use" of any substance defined as “(reported ≥1 standard alcohol drink, >puff/taste cannabis or nicotine or any other occasion of consumption/use)” (Sullivan et al, 2022). Our implemented SU definition operationalizes this with the addition of objective toxicology, which was not specified at pre-registration but increases measurement validity due to our observed discordance (replicating Wade et al, 2023; Supplementary Table 6).

Four alternative SU operationalizations were considered:

1. TLFB/iSIP self-report alone (no toxicology). We replicated prior literature documenting low concordance between adolescent self-report and toxicology (Supplementary Table 6; Wade et al, 2023), which would have introduced systematic misclassification rather than reflecting a methodological decision. Therefore, our analyses incorporated toxicology data (see Supplementary Methods, section 3 and Supplementary Table 6).
2. Substance-specific outcomes (e.g., cannabis-only or alcohol-only models): not pursued because polysubstance use was common (Supplementary Table 7).
3. Wave-specific definitions were examined (use at Y4 only or Y6 only), but were not adopted as primary because they a) yielded co-occurrence group sizes (Y4-only: n = 88) below the minimum n = 100 per-group threshold specified in the pre-registration as required for adequately powered comparisons, and b) discarded information by ignoring use documented at the other wave.

4. Stricter "more than one day of use" definition. A stricter iteration, requiring more than a single "full use" of any substance at Y4 or Y6 was also considered. Preliminary multi-seed comparison testing suggested modestly improved SU-only vs. ON prediction under this definition, reflecting sharper discrimination from excluding one-time experimenters, but at the cost of reducing the co-occurrence group by ~38% (n = 103). Given that the co-occurrence group is a foundational comparison for our research questions, this reduction was judged to outweigh the marginal predictive benefit and the definition was not adopted for the primary analyses.

These considerations supported the "full-use" definition, in line with our preregistered intent. All findings reported reflect this definition.

Outcome group assignment. Based on the PLE and full-use SU classifications, participants were assigned to one of four mutually exclusive groups:

- Outcome-negative (ON): did not meet threshold for PLE at any wave and did not report or screen positive for SU at any wave (n = 5,824)
- PLE-only: met PLE threshold at one or more waves but did not meet SU criteria (n = 281)
- SU-only: met SU criteria at one or more waves but did not meet PLE threshold (n = 3,703)
- Co-occurrence: met both PLE and SU criteria across the relevant waves (n = 326)

**Supplementary Methods 3:** Toxicology Measures (Hair, Urine, Salivary, and Breathalyzer)**.**

We incorporated toxicology data from year Y4 and Y6 biospecimen screenings at follow-up visits into the SU outcome definition in order to complement the self-report substance use from the TLFB interview and iSIP screener. Samples were processed by the ABCD Study's centralized laboratory (methodology previously described) and returned as indicators by substance class. We analyzed these indicators as binary yes/no.

Hair samples included screenings cannabinoids, cocaine, opiates, amphetamines, fentanyl, nicotine, alcohol, and other commonly used substances. Urine samples were screened using standard immunoassay panels for recent substance and included THC, cocaine metabolites, opiates, nicotine, amphetamines, benzodiazepines, and other substances. Oral fluid and salivary samples were collected at annual visits and screened for THC, amphetamines, cocaine, methamphetamine, opioids, methadone, and benzodiazepines, and nicotine. Breath alcohol concentration was measured at in-person visits using a handheld breathalyzer device.

A participant was classified as having used substances at a given follow-up wave if they endorsed substance use on the TLFB interview and/or the iSIP screener, and/or screened positive on any toxicology measure. The final SU outcome reflected meeting this criterion at either the Y4 or Y6 follow-up. Participants with missing toxicology data at a given wave were classified based on available TLFB and iSIP screener data alone, and participants missing both self-report and toxicology at both Y4 and Y6 were treated as attrition cases and excluded from analyses (see Outcome Group Construction, Attrition and Supplementary Figure 1: Sample construction flow chart).

**Counts of toxicology-positive participants by substance and wave are presented in Supplementary Table 6.**

**A complete inventory of all self-report and toxicology variables used to construct the substance use outcome including substance category, source instrument, ABCD variable name, label, time point, and role in outcome derivation is provided in Supplementary Table 5 (provided as a separate file: SUPP_TABLE5_substance_use_variables.csv).**

**Supplementary Methods 4. Baseline input features**

A complete inventory of all baseline features used in the prediction models, including domain, variable name, ABCD source variable(s), derivation method, and description, is provided in Supplementary Table 4 (provided as a separate file: SUPP_TABLE4_baseline_features.csv).

**Supplementary Methods 5: Preprocessing Pipeline**

Parameters were estimated exclusively from training folds and applied to held-out test data. All preprocessing steps were performed within the cross-validation loop.

Pre-registered nuisance covariates were regressed out of imaging features prior to model training. For each feature, an ordinary least squares regression was fit on training-fold data. Site effects, scanner manufacturer, and scanner serial number were included as covariates. For functional imaging features (resting-state functional connectivity and task-based activation), modality-specific mean framewise displacement was additionally included as a covariate to account for residual motion-related variance. Fitted regression coefficients from the training fold were then applied to the held-out test fold to produce residualized test-set features. For ridge, elastic net, SVC, and random forest models, continuous features were standardized to zero mean and unit variance within training folds, with the same transformation applied to held-out test folds using training-fold parameters.

XGBoost learns a default branch direction for missing values at each split allowing the model to incorporate informative missingness patterns without imposing imputation assumptions. Thus missing values were handled natively during tree construction for XGBoost models. Missing values were imputed within training folds suing median imputation for continuous features and mode imputation for categorical features. The same training-fold imputation values were applied to test folds.

Within each cross-validation fold, we applied preprocessing in the following order: 1) stratified train-test split, 2) residualization of imaging features on nuisance covariates, 3) missing data handling (imputation for comparison models; native handling for XGBoost), 4) standardization of continuous for ridge, elastic net, SVC, random forest.

**Supplementary Methods 6:** Model specification and hyperparameters.

Five supervised classification algorithms were evaluated in accordance with the pre-registered analysis plan, including ridge, elastic net, support vector classifiers (SVC), random forest, and extreme gradient boosting (XGBoost). Each model was trained on three feature configurations: combined features, neuroimaging features only, and non-neuroimaging features only, for each of the six binary outcome comparisons. Performance across all models, feature sets, and comparisons is reported in Supplementary Table 2.

**Rationale for Binary Outcome versus Outcome-Negative Classification Approach.** Rather than treating the four outcome groups (PLE-only, SU-only, co-occurrence, ON) as a single multiclass classification problem, we decomposed the analysis into a series of binary comparisons due to analytic design considerations. First, multiclass classification with severe class imbalance across groups produces classifiers that are dominated by the majority class and yield uninterpretable performance metrics. In our case, the ON group (n=5,824) outnumbered the smallest outcome group (PLE-only, n=281) by more than 20:1. Our binary comparison approach allowed us to address imbalance separately for each outcome via targeted downsampling and scale_pos_weight calibration. Second, the scientific questions motivating each comparison are fundamentally distinct: predicting who will develop persistent PLEs versus predicting who will initiate substance use call on different feature sets, and therefore have different expected predictability. Collapsing them into a single multiclass model would obscure these differences. Third, binary classification yields directly interpretable AUROC values that are comparable across outcomes, and more comparable to existing literature, whereas multiclass metrics are harder to interpret and less commonly reported in clinical prediction research. This approach allowed us to contextualize our classifications within the broader literature and leant to interpretability. Fourth, the binary framework allows for targeted SHAP-based feature importance analysis within each comparison. This approach enables us to identify which features specifically drive discrimination for each outcome, rather than averaging across heterogeneous outcome classes. In summary, the binary comparisons reported in our analyses collectively address the same scientific questions that a multiclass model would, while providing more granular, and more interpretable results.

All models were implemented in Python using scikit-learn. XGBoost was performed with the xgboost library. Class imbalance was addressed using class_weight='balanced' for the comparison models and scale_pos_weight for XGBoost, with the modeling pipeline additionally applying controlled downsampling of the majority class where appropriate (see Outcome Group Construction and Attrition). Hyperparameters were tuned via stratified 5-fold cross-validation within training data, with final performance evaluated on held-out data. All models were evaluated using identical feature sets and identical train-test partitions within each comparison to enable direct performance comparison.

Ridge and elastic net classification were implemented using scikit-learn's LogisticRegression with L2 and elastic net penalties, respectively. For elastic net, both the regularization strength (C) and the L1/L2 mixing parameter (l1_ratio) were tuned. SVC was implemented using SVC with probability estimates enabled. Random forest was implemented using RandomForestClassifier, with tuning over the number of estimators, maximum tree depth, and minimum samples per split and leaf. For all four comparison models, hyperparameters were selected via nested stratified 5-fold cross-validation using out-of-fold predictions, with preprocessing wrapped in scikit-learn pipelines to prevent leakage. Best-performing hyperparameter values are provided in Supplementary Table 9.

XGBoost was implemented using XGBClassifier with the histogram tree method (tree_method='hist'), logistic binary objective, and AUROC as the evaluation metric. Early stopping (50 rounds) was applied using held-out test data to prevent overfitting. Hyperparameter tuning was conducted using Bayesian optimization (BayesSearchCV from scikit-optimize) over 75 iterations, with a fallback to randomized search if Bayesian optimization failed. The search space included:

- max_depth: integer, 2–10
- learning_rate: log-uniform, 0.001–0.3
- subsample: uniform, 0.5–1.0
- colsample_bytree: uniform, 0.5–1.0
- colsample_bylevel: uniform, 0.5–1.0
- min_child_weight: integer, 1–20
- reg_alpha: log-uniform, 1e-8 to 1.0
- reg_lambda: log-uniform, 1e-6 to 10.0
- n_estimators: integer, 100–5000

XGBoost was evaluated using repeated stratified 80/20 train-test splits (5 iterations), with inner stratified 5-fold cross-validation for hyperparameter tuning within each training split. Performance is reported as the mean test AUROC ± standard deviation across splits, with range. Interpretability analyses (SHAP values) were derived from the highest-performing split, and aggregated SHAP values across all 5 splits are additionally reported (see SHAP Value Computation and Interpretation). Best-performing XGBoost hyperparameters for each comparison and feature set are provided in Supplementary Table 9.

**Supplementary Methods 7.** Validation and Sensitivity Analyses

We conducted a series of complementary model validation and sensitivity analyses for all XGBoost classification analyses to validate that observed predictive performance reflected meaningful signal rather than data leakage, model overfitting, or spurious associations.

Cross-validation and leakage control

All models were trained and evaluated using a nested cross-validation framework, with preprocessing steps-including covariate residualization, scaling (for linear and distance-based comparison models), and imputation (for comparison models requiring complete feature matrices; XGBoost handled missingness natively), performed exclusively within training folds and applied to held-out test data. Model performance was evaluated on held-out data only, and results were aggregated across cross-validation folds to assess stability.

Label permutation testing

To further assess whether model performance reflected true structure in the data rather than chance correlations or pipeline artifacts, label permutation analyses were conducted. Outcome labels were randomly permuted while preserving the original feature structure, and the full modeling pipeline was re-run on permuted data. Under label permutation, model performance consistently collapsed to near-chance levels, supporting the absence of information leakage and confirming that observed accuracy depended on the correct alignment between features and outcomes (Supplementary Table 3).

Feature set ablation analyses

Model performance was additionally evaluated across multiple feature configurations, including models trained on neuroimaging features alone, non-neuroimaging features alone, and combined feature sets.

Stability across random seeds and folds

Finally, robustness was assessed across multiple random seeds and cross-validation splits. Performance metrics were consistent across folds, indicating that results were not driven by a single favorable data partition or random initialization.

Use of baseline PLE input feature

As a sensitivity analysis, we re-ran our models with baseline PLEs excluded as an input feature to test whether model performance could be attributed to their inclusion. We found that model performance slightly dropped across models: PLE-only vs. ON AUROC = 0.713 (vs. 0.780 in the primary analysis), SU-only vs. ON AUROC = 0.623 (vs. 0.641), and Co-occurrence vs. ON AUROC = 0.775 (vs. 0.828). Differences were modest (mean $|\Delta| = 0.041$), comparable in magnitude to per-comparison fold variability (mean SD = 0.027), indicating that the predictive signal in our primary models was not driven by baseline PLE inclusion.

**Supplementary Methods 8:** SHAP Value Computation and Interpretation.

We computed SHAP (SHapley Additive exPlanations) values for each model in order to interpret model predictions. This method decomposes a model's prediction for each individual into additive contributions from each input feature. SHAP values were computed using the TreeSHAP algorithm shap.TreeExplainer, which utilizes the structure of tree-based models to compute exact Shapley values in polynomial time. SHAP values were calculated on held-out test data for each outer train-test split, using a random subsample of up to 3,000 test-set observations per split. The sum of SHAP values across all features, plus a baseline expected value, equals the model's predicted output for a given individual. For binary classification with XGBoost, SHAP values are expressed in the model's margin space (log-odds). A SHAP value for a given feature quantifies that feature's additive contribution to the predicted log-odds of the positive class for a given individual, relative to the model's expected (baseline) prediction. Positive SHAP values indicate that a feature's value pushes the prediction toward the positive class, while negative SHAP values indicate that the feature's value pushes the prediction toward the negative class.

To summarize the overall importance of each feature within a given model, we computed the mean absolute SHAP value across individuals for each feature. This quantity reflects the average magnitude of a feature's contribution to predictions regardless of direction. We computed a directional SHAP summary by multiplying each feature's mean absolute SHAP value by the sign of the Spearman correlation between that feature's values and its SHAP values across individuals to additionally capture the direction of each feature's typical contribution. This yields a signed importance measure where positive values indicate that higher feature values tend to increase the predicted probability of the positive class, and negative values indicate that higher feature values tend to decrease it. This provides an interpretable complement to the undirected mean absolute SHAP metric.

SHAP values were computed separately within each split. To summarize feature importance across splits, we computed the mean and standard deviation of mean absolute SHAP values across the 5 splits, as well as the mean directional SHAP value across splits. Aggregated SHAP values across splits are reported alongside the single-split SHAP values from the highest-performing split (which are visualized in the main text and in Supplementary Figures 3-4). SHAP beeswarm plots (Supplementary Figure 3) display the distribution of SHAP values for each of the top-ranked features, with individual observations shown as points colored by feature value. Complementary bar plots of mean absolute SHAP values and directional SHAP summaries are provided in Supplementary Figure 4.

**Supplementary Table 1:** Participant Demographics by Outcome Group

| Variable | Outcome-negative (n = 5,823) | Psychotic-like Experiences (n = 281) | Substance Use (n = 3,703) | Co-occurring PLEs + SU (n = 326) | *p* |
|---|---|---|---|---|---|
| Age, mean (SD) | 9.91 (0.62)[bc] | 9.72 (0.58)[acd] | 10.04 (0.62)[abd] | 9.93 (0.62)[bc] | <0.001 |
| Female, N (%) | 2647 (45.5)[bcd] | 172 (61.2)[ac] | 1771 (47.8)[abd] | 222 (68.1)[ac] | <0.001 |
| $0-24,999 | 659 (12.3)[bcd] | 53 (20.5)[ac] | 510 (14.9)[abd] | 60 (20.8)[ac] | <0.001 |
| $25k-49,999 | 671 (12.5)[bcd] | 70 (27.0)[ac] | 517 (15.1)[abd] | 74 (25.7)[ac] | <0.001 |
| $50k-74,999 | 749 (13.9) | 37 (14.3) | 488 (14.2) | 37 (12.8) | 0.917 |
| $75-$99,999 | 819 (15.2) | 37 (14.3) | 493 (14.4) | 33 (11.5) | 0.273 |
| $100k-$199,999 | 1825 (34.0)[bcd] | 53 (20.5)[ac] | 999 (29.1)[abd] | 65 (22.6)[ac] | <0.001 |
| $200k or more | 652 (12.1)[bd] | 9 (3.5)[ac] | 421 (12.3)[bd] | 19 (6.6)[ac] | <0.001 |
| White, N (%) | 4367 (75.2)[bcd] | 168 (59.8)[ac] | 2906 (78.6)[abd] | 213 (65.5)[ac] | <0.001 |
| Black, N (%) | 1153 (19.9)[bd] | 101 (35.9)[ac] | 711 (19.2)[bd] | 99 (30.5)[ac] | <0.001 |
| Asian, N (%) | 429 (7.4)[c] | 12 (4.3) | 181 (4.9)[a] | 16 (4.9) | <0.001 |
| Pacific Islander, N (%) | 43 (0.7) | 0 (0.0) | 20 (0.5) | 1 (0.3) | 0.274 |
| Native American, N (%) | 156 (2.7)[bc] | 14 (5.0)[a] | 163 (4.4)[a] | 13 (4.0) | <0.001 |
| Other, N (%) | 362 (6.2)[bd] | 33 (11.7)[ac] | 223 (6.0)[bd] | 31 (9.5)[ac] | <0.001 |
| Hispanic, N (%) | 1010 (17.4)[bd] | 71 (25.3)[ac] | 693 (18.8)[b] | 74 (22.8)[a] | <0.001 |

Legend:
a = Outcome-negative (ON)
b = Psychotic-like Experiences
c = Substance Use
d = Co-occurring PLEs + SU

Baseline demographics by outcome group. Continuous variables are reported as mean (SD); categorical variables as N (%). Group differences were tested via one-way ANOVA (continuous) or chi-square (categorical), with pairwise follow-up tests (Welch's t-tests or pairwise chi-square). Superscripts denote significant pairwise differences ($p < 0.05$). Race categories are not mutually exclusive; participants who endorsed more than one race are included in each applicable category.

**Supplementary Table 2:** Per-Fold and Per-Seed AUROC Values

| Comparison | CV AUROC | Test AUROC | Brier Score | Sensitivity | Specificity | PPV | NPV |
|---|---|---|---|---|---|---|---|
| PLE vs. ON | 0.793 ± 0.013 | 0.780 ± 0.050 | 0.186 ± 0.020 | 0.614 ± 0.112 | 0.800 ± 0.026 | 0.378 ± 0.058 | 0.913 ± 0.024 |
| SU vs. ON | 0.623 ± 0.002 | 0.626 ± 0.007 | 0.240 ± 0.005 | 0.557 ± 0.023 | 0.626 ± 0.017 | 0.487 ± 0.010 | 0.690 ± 0.009 |
| Co-occurrence vs. ON | 0.793 ± 0.008 | 0.828 ± 0.023 | 0.194 ± 0.043 | 0.661 ± 0.070 | 0.821 ± 0.059 | 0.434 ± 0.066 | 0.925 ± 0.012 |
| Co-occurrence vs. SU | 0.735 ± 0.004 | 0.765 ± 0.030 | 0.228 ± 0.020 | 0.637 ± 0.090 | 0.740 ± 0.015 | 0.176 ± 0.015 | 0.959 ± 0.009 |
| Co-occurrence vs. PLE | 0.565 ± 0.015 | 0.596 ± 0.034 | 0.250 ± 0.008 | 0.612 ± 0.078 | 0.543 ± 0.086 | 0.614 ± 0.019 | 0.544 ± 0.017 |
| PLE vs. SU | 0.753 ± 0.007 | 0.771 ± 0.026 | 0.211 ± 0.033 | 0.511 ± 0.224 | 0.812 ± 0.105 | 0.194 ± 0.061 | 0.958 ± 0.013 |

Supplementary Table 2a. XGBoost performance across all metrics, reported as mean ± standard deviation across 10 stratified train-test splits. Brier score reflects probabilistic calibration (lower is better); sensitivity, specificity, PPV, and NPV are computed at a 0.5 classification threshold.

| Comparison | Feature Set | XGBoost | Elastic Net | Ridge | Random Forest | SVC |
|---|---|---|---|---|---|---|
| PLE vs. ON | All | 0.780 ± 0.050 | 0.781 ± 0.037 | 0.772 ± 0.036 | 0.771 ± 0.036 | 0.743 ± 0.042 |
| PLE vs. ON | Non-Neuroimaging-only | 0.783 ± 0.038 | 0.785 ± 0.040 | 0.784 ± 0.040 | 0.788 ± 0.035 | 0.745 ± 0.048 |
| PLE vs. ON | Neuroimaging-only | 0.571 ± 0.024 | 0.521 ± 0.007 | 0.518 ± 0.020 | 0.534 ± 0.022 | 0.535 ± 0.035 |
| SU vs. ON | All | 0.626 ± 0.007 | 0.618 ± 0.006 | 0.618 ± 0.006 | 0.620 ± 0.008 | 0.615 ± 0.004 |
| SU vs. ON | Non-Neuroimaging-only | 0.627 ± 0.010 | 0.619 ± 0.005 | 0.620 ± 0.005 | 0.619 ± 0.006 | 0.617 ± 0.004 |
| SU vs. ON | Neuroimaging-only | 0.543 ± 0.010 | 0.518 ± 0.012 | 0.516 ± 0.011 | 0.525 ± 0.014 | 0.513 ± 0.013 |
| Co-occurrence vs. ON | All | 0.828 ± 0.023 | 0.826 ± 0.025 | 0.818 ± 0.021 | 0.823 ± 0.022 | 0.790 ± 0.020 |
| Co-occurrence vs. ON | Non-Neuroimaging-only | 0.835 ± 0.028 | 0.826 ± 0.025 | 0.827 ± 0.023 | 0.829 ± 0.027 | 0.796 ± 0.025 |
| Co-occurrence vs. ON | Neuroimaging-only | 0.615 ± 0.018 | 0.599 ± 0.033 | 0.566 ± 0.033 | 0.587 ± 0.024 | 0.585 ± 0.022 |
| Co-occurrence vs. SU | All | 0.765 ± 0.030 | 0.776 ± 0.022 | 0.765 ± 0.026 | 0.734 ± 0.030 | 0.745 ± 0.028 |
| Co-occurrence vs. SU | Non-Neuroimaging-only | 0.764 ± 0.023 | 0.778 ± 0.020 | 0.771 ± 0.023 | 0.751 ± 0.023 | 0.749 ± 0.017 |
| Co-occurrence vs. SU | Neuroimaging-only | 0.572 ± 0.019 | 0.563 ± 0.009 | 0.549 ± 0.015 | 0.512 ± 0.024 | 0.556 ± 0.033 |
| Co-occurrence vs. PLE | All | 0.596 ± 0.034 | 0.564 ± 0.046 | 0.572 ± 0.048 | 0.554 ± 0.035 | 0.544 ± 0.081 |
| Co-occurrence vs. PLE | Non-Neuroimaging-only | 0.623 ± 0.047 | 0.607 ± 0.042 | 0.611 ± 0.043 | 0.574 ± 0.044 | 0.422 ± 0.082 |
| Co-occurrence vs. PLE | Neuroimaging-only | 0.557 ± 0.021 | 0.472 ± 0.060 | 0.462 ± 0.050 | 0.509 ± 0.032 | 0.492 ± 0.032 |
| PLE vs. SU | All | 0.771 ± 0.026 | 0.786 ± 0.011 | 0.777 ± 0.012 | 0.757 ± 0.026 | 0.764 ± 0.018 |
| PLE vs. SU | Non-Neuroimaging-only | 0.769 ± 0.020 | 0.780 ± 0.011 | 0.773 ± 0.012 | 0.759 ± 0.016 | 0.764 ± 0.010 |
| PLE vs. SU | Neuroimaging-only | 0.575 ± 0.040 | 0.597 ± 0.026 | 0.593 ± 0.024 | 0.542 ± 0.022 | 0.530 ± 0.049 |

Supplementary Table 2b. Test AUROC (mean ± standard deviation) across outer cross-validation folds for each binary classification task and feature set. XGBoost achieved the highest or tied-highest mean AUROC in 11 of 18 analyses, with an advantage on neuroimaging-feature models (mean advantage +0.021 AUROC over elastic net), consistent with nonlinear and interactive structure in neuroimaging predictors. On all-feature and non-neuroimaging-only models, performance differences between XGBoost and linear comparators were generally within fold-to-fold variability. ON = outcome-negative comparison group.

**Supplementary Table 3**. Negative control analysis using random labels

We conducted a negative control analysis by replacing the true outcome labels with randomly generated binary labels across all three primary comparisons to evaluate whether model performance could arise from unintended data leakage or biases in the modeling pipeline. The full modeling pipeline, including preprocessing, residualization, feature selection, hyperparameter tuning, and model fitting was run without modification.

As expected under the absence of a true signal, model performance was at or near chance levels across all three comparisons. Cross-validated AUROC ranged from 0.505 to 0.542, and test AUROC ranged from 0.459 to 0.547, with accuracy approximating 0.50 in all cases. Precision, recall, and F1-scores were similarly balanced across classes.

| Comparison | CV AUROC | Test AUROC | Accuracy | Class 0 F1 | Class 1 F1 | Macro F1 |
|---|---|---|---|---|---|---|
| PLE vs. ON | 0.542 | 0.459 | 0.509 | 0.0 | 0.67 | 0.34 |
| SU vs. ON | 0.505 | 0.507 | 0.511 | 0.67 | 0.09 | 0.38 |
| Co-occurrence vs. ON | 0.521 | 0.547 | 0.528 | 0.26 | 0.65 | 0.46 |

**Supplementary Table 4:** Full List of Baseline Input Features

A complete inventory of all baseline features used in the prediction models, including domain, variable name, ABCD source variable(s), derivation method, and description, is provided in Supplementary Table 4 (provided as a separate file: SUPP_TABLE4_baseline_features.csv).

**Supplementary Table 5:** Summary of substance use measures and variables

A complete inventory of all self-report and toxicology variables used to construct the substance use outcome including substance category, source instrument, ABCD variable name, label, time point, and role in outcome derivation is provided in Supplementary Table 5 (provided as a separate file: SUPP_TABLE5_substance_use_variables.csv)

**Supplementary Table 6**: Self-Report vs Toxicology Concordance

Self-report vs. toxicology Year 4:

| Self-Report | Tox Negative | Tox Positive |
|---|---|---|
| No Use Reported (0) | 7,634 | 1,258 |
| Use Reported (1) | 576 | 267 |

False Negative Rate: among those reporting no use, 14.15% tested positive.
False Positive Rate: among those reporting use, 7.02% tested negative.
Positive Predictive Value: among those reporting use, 31.7% were toxicology-confirmed

Self-report vs. toxicology Year 6:

| Self-Report | Tox Negative | Tox Positive |
|---|---|---|
| No Use Reported (0) | 2,883 | 751 |
| Use Reported (1) | 758 | 664 |

False Negative Rate: among those reporting no use, 20.67% tested positive.
False Positive Rate: among those reporting use, 20.82% tested negative.
Positive Predictive Value: among those reporting use, 46.7% were toxicology-confirmed

Concordance between self-reported substance use (TLFB and iSIP) and toxicology screen results at Year 4 (panel a) and Year 6 (panel b), restricted to participants in the analytic sample (*N* = 10,134) with both data sources available at each wave (n = 9,735 at Y4; n = 5,056 at Y6). Among participants reporting no use on TLFB or iSIP, 14.15% tested positive on toxicology at Y4 and 20.67% tested positive at Y6 (false negative rates; false positive rates and positive predictive value reported above), a pattern consistent with the literature on increasing underreporting of substance use as adolescents age (e.g. Wade et al, 2023). The discordance at both waves motivated the inclusion of toxicology data in the substance use outcome definition (see Supplementary Methods 2).

**Supplementary Table 7:** Rates of polysubstance use

| Wave | Mono-substance users | Polysubstance users (≥2 substances) | Positive for all three substances |
|---|---|---|---|
| Y4 | 72.4% | 27.6% | 7.4% |
| Y6 | 55.0% | 45.0% | 17.2% |

This study was underpowered to detect substance-specific effects as polysubstance use was common and increased substantially with age: among Y4 users, 27.6% endorsed two or more substances, rising to 45.0% by Y6, with 321 participants reporting use of all three substances at Y6. Substance use was also moderately intercorrelated at Y6 (alcohol–nicotine $\varphi = 0.39$, alcohol–cannabis $\varphi = 0.33$, nicotine–cannabis $\varphi = 0.45$).

Substance use rates at Year 4 and Year 6 follow-up as assessed via toxicology, summarized across toxicology measures. Participants were classified as positive for a given substance if they reported any use on the TLFB, iSIP, or tested positive on toxicology (urine, saliva, hair, and/or alcohol breathalyzer, depending on the substance). "All Substances" reflects the union of use across all substances.

**Supplementary Table 8**: Sample Sizes and Downsampling Parameters by Comparison

| Comparison | Positive class | Negative class | N positive | N negative (available) | N negative (used) | Downsampling ratio | Total N (analytic) | % positive | scale_pos_weight |
|---|---|---|---|---|---|---|---|---|---|
| PLE-only vs. ON | PLE only | ON | 281 | 5,823 | 1,405 | 5:1 (PLE_only:negatives) | 1,686 | 16.7% | 5.00 |
| SU-only vs. ON | SU only | ON | 3,703 | 5,823 | 5,823 | ON (all available negatives used) | 9,526 | 38.9% | 1.57 |
| Co-occurrence vs. ON | Co-occurrence | ON | 326 | 5,823 | 1,630 | 5:1 (both:negatives) | 1,956 | 16.7% | 5.00 |
| Co-occurrence vs. SU-only | Co-occurrence | SU only | 326 | 3,703 | 3,703 | ON (all available negatives used) | 4,029 | 8.1% | 11.36 |
| Co-occurrence vs. PLE-only | Co-occurrence | PLE only | 326 | 281 | 281 | ON (all available negatives used) | 607 | 53.7% | 0.86 |
| PLE-only vs. SU-only | PLE only | SU only | 281 | 3,703 | 3,703 | ON (all available negatives used) | 3,984 | 7.1% | 13.18 |

Sample sizes, downsampling parameters, and scale_pos_weight values for each binary comparison in the analytic sample (N = 10,134). For comparisons against the ON group, the negative pool was downsampled to a 5:1 negative-to-positive ratio for the rare-positive comparisons (PLE-only vs. ON and Co-occurrence vs. ON) to balance computational efficiency with adequate signal preservation, while the SU-only vs. ON comparison used all available negatives, given its more balanced base rate. Secondary comparisons (Co-occurrence vs. SU-only, Co-occurrence vs. PLE-only, PLE-only vs. SU-only) used all available participants in both classes without downsampling. The scale_pos_weight parameter was set to the negative-to-positive ratio of the training set and was used by XGBoost to upweight minority-class errors during gradient boosting; for comparison models (random forest, ridge regression, elastic net, support vector classifier), an equivalent class_weight = "balanced" adjustment was applied. Class imbalance varied substantially across comparisons, ranging from 7.1% positive (PLE-only vs. SU-only) to 53.7% positive (Co-occurrence vs. PLE-only); this variation was directly handled by the per-comparison weighting scheme rather than by oversampling, given prior evidence that synthetic oversampling (e.g., SMOTEENN) introduced substantial overfitting in our pilot analyses.

**Supplementary Table 9:** XGBoost and Comparison Model Hyperparameters

| Comparison | Max Depth | Learning Rate | N Estimators | Subsample | Col/Tree | Col/Level | Min Child Weight | reg_alpha | reg_lambda |
|---|---|---|---|---|---|---|---|---|---|
| PLE vs. ON | 7 | 0.077 | 4607 | 0.992 | 0.737 | 0.596 | 20 | <0.001 | 0.015 |
| SU vs. ON | 6 | 0.006 | 1298 | 0.598 | 0.742 | 0.686 | 14 | 0.017 | 0.004 |
| Co-occurrence vs. ON | 5 | 0.005 | 2194 | 0.522 | 0.764 | 0.834 | 2 | <0.001 | 0.003 |
| Co-occurrence vs. SU | 2 | 0.013 | 137 | 0.706 | 0.541 | 0.967 | 14 | <0.001 | <0.001 |
| Co-occurrence vs. PLE | 2 | 0.053 | 3632 | 0.922 | 0.605 | 0.531 | 9 | 0.350 | 0.139 |
| PLE vs. SU | 2 | 0.013 | 137 | 0.706 | 0.541 | 0.967 | 14 | <0.001 | <0.001 |

Supplementary Table 9a. XGBoost best-performing hyperparameters by comparison (all features, best cross-validation split). Col/Tree = colsample_bytree (fraction of features sampled per tree); Col/Level = colsample_bylevel (fraction of features sampled per tree level); reg_alpha = L1 regularization term; reg_lambda = L2 regularization term. Gamma = 0 for all comparisons. Hyperparameters reflect the best-performing outer cross-validation split identified via Bayesian optimization over 75 iterations, with random search as a fallback. Early stopping was applied with a patience of 50 rounds using held-out test data.

| Model | Hyperparameter | Search Values | Final Setting |
|---|---|---|---|
| Ridge | C (regularization) | 0.01, 0.1, 1.0, 10.0 | Best via 5-fold CV |
| Elastic Net | C (regularization) | 0.01, 0.1, 1.0 | Best via 5-fold CV |
| Elastic Net | l1_ratio | 0.3, 0.5, 0.7 | Best via 5-fold CV |
| SVC | C (regularization) | 0.1, 1.0, 10.0 | Best via 5-fold CV |
| Random Forest | n_estimators | 500 (fixed) | 500 |
| Random Forest | max_depth | 10 (fixed) | 10 |
| Random Forest | min_samples_leaf | 5 (fixed) | 5 |

Supplementary Table 9b. Hyperparameter search grids for comparison models. All comparison models used stratified cross-validation for hyperparameter selection via grid search.

**Supplementary Figure 1:** Sample construction flow chart

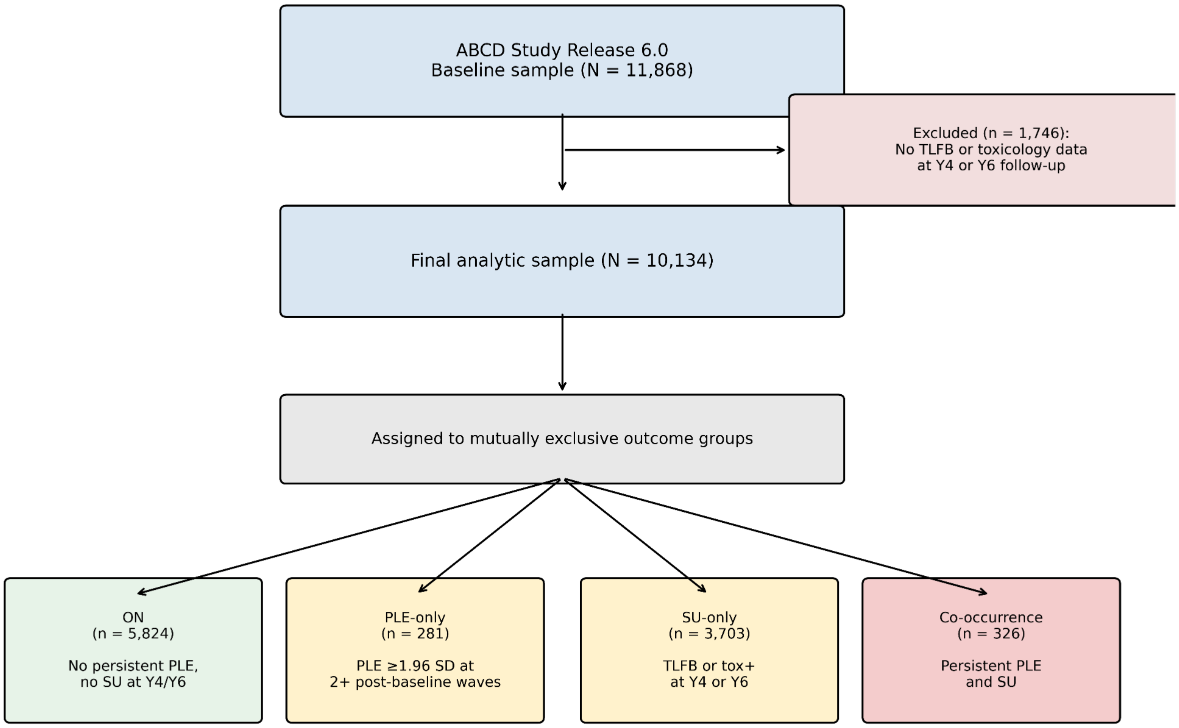


Sample construction flow chart. Of the 11,868 participants in the ABCD Study Release 6.0 baseline sample, 1,746 (14.71%) were excluded because they had no available substance use data (no Timeline Followback self-report and no toxicology screen) at either the Y4 or the Y6 follow-up wave, yielding a final analytic sample of N = 10,134. Participants were assigned to one of four mutually exclusive outcome groups based on persistent psychotic-like experiences (PQ-BC distress ≥1.96 SD above the wave-specific mean at two or more annual post-baseline assessment waves, Years 1 through 6) and substance use (self-reported use or positive toxicology at Y4 or Y6): ON (n = 5,824), PLE-only (n = 281), SU-only (n = 3,703), and Co-occurrence (n = 326). Within the analytic sample, imaging features were set to missing on a per-participant basis when task-fMRI behavioral quality control failed or when resting-state functional MRI mean framewise displacement exceeded 0.9 mm; these participants remained in the analysis, and XGBoost handled missing imaging values natively during tree construction without requiring imputation

**Supplementary Figure 2:** Model Performance Stability Across Cross-Validation Folds and Random Seeds

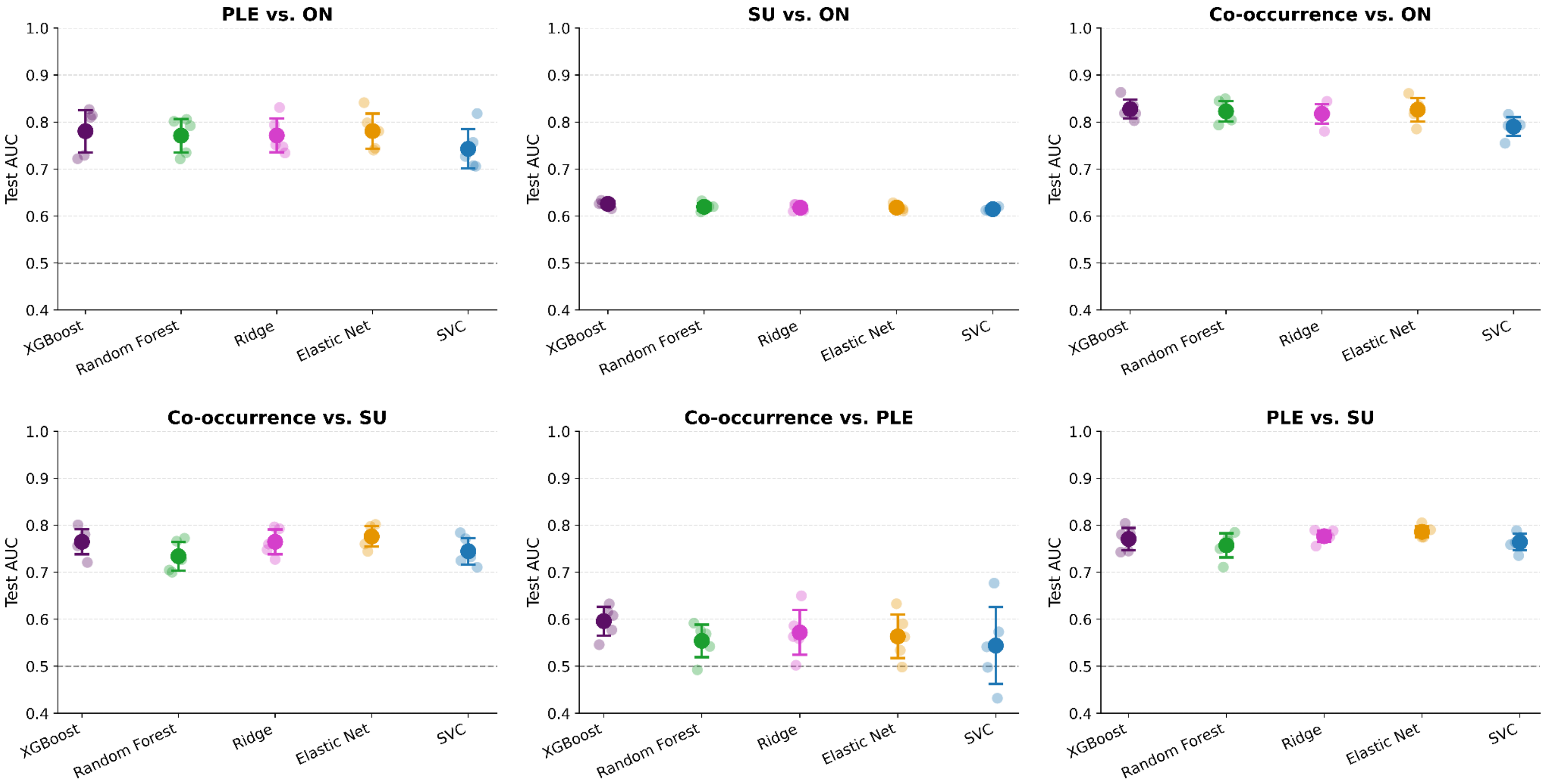


Test AUROC is shown for each of five model types (XGBoost, Random Forest, Ridge logistic regression, Elastic Net, and Support Vector Classifier) across the three primary binary comparisons using the full feature set (neuroimaging + non-neuroimaging combined). Individual points represent per-fold (XGBoost) or per-seed (all other models) AUROC values, with larger markers and error bars indicating mean ± one standard deviation. For XGBoost, variability is estimated from 5 stratified outer cross-validation splits. For all other models, variability reflects 5 random seeds. The dashed horizontal line indicates chance performance (AUROC = 0.50). The tight clustering of individual points around the mean demonstrates consistent performance across evaluation splits and seeds, supporting the robustness of the reported findings.

**Supplementary Figure 3:** Precision-Recall Curves

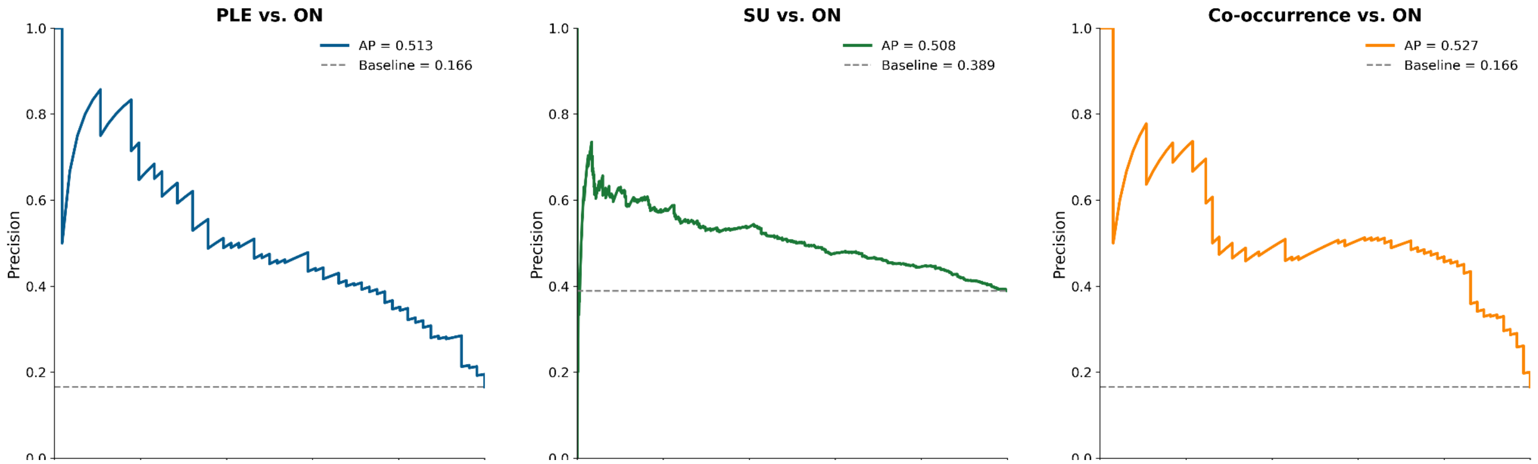


Precision-recall curves for the three primary binary comparisons (PLE vs. ON, SU vs. ON, co-occurrence vs. ON) using XGBoost with the full feature set. The dashed horizontal line represents the no-skill baseline equal to the prevalence of the positive class in each comparison. Average precision (AP) is reported for each comparison. Given the class imbalance inherent to these comparisons, precision-recall curves provide a complementary and more sensitive characterization of model performance than ROC curves alone. Precision-recall curves are particularly informative under class imbalance, as average precision is benchmarked against the prevalence baseline rather than a fixed chance level (Saito & Rehmsmeier, 2015); modest absolute gains in precision are therefore expected in this low-base-rate setting.

**Supplementary Figure 4:** Calibration Plots Across Comparisons

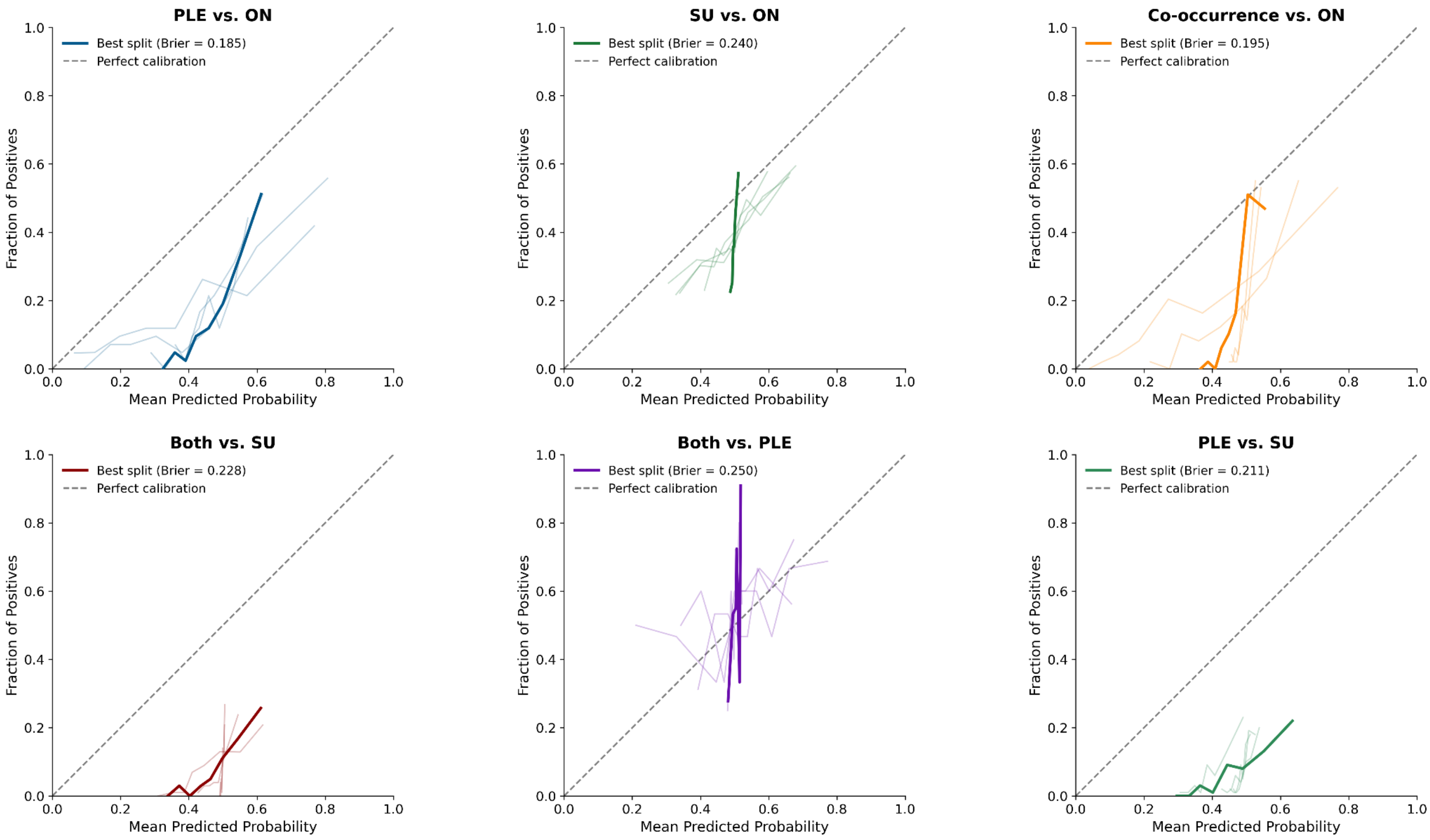


Calibration curves for the XGBoost model across all six binary comparisons using the full feature set. Each panel shows calibration curves for all 5 cross-validation splits (faint lines) and the best-performing split (bold line). The diagonal dashed line represents perfect calibration. The mean Brier score across splits is reported in the legend for each comparison. Deviations above the diagonal indicate underconfidence and deviations below indicate overconfidence. Given the class imbalance in these comparisons, predicted probabilities are generally low; however, the curves demonstrate that the model's relative probability ordering is meaningful and consistent across splits. Tree-based ensembles XGBoost can produce probability estimates not natively well-calibrated under class imbalance. Observed deviations from the diagonal are therefore expected and reflect a known property of XGBoost rather than a failure of the underlying classifier. AUROC and rank ordering are not affected by these calibration patterns and predicted probabilities could be recalibrated post-hoc, e.g., via Platt scaling or isotonic regression.

**Supplementary Figure 5**: Directional feature importance for PLEs and SU

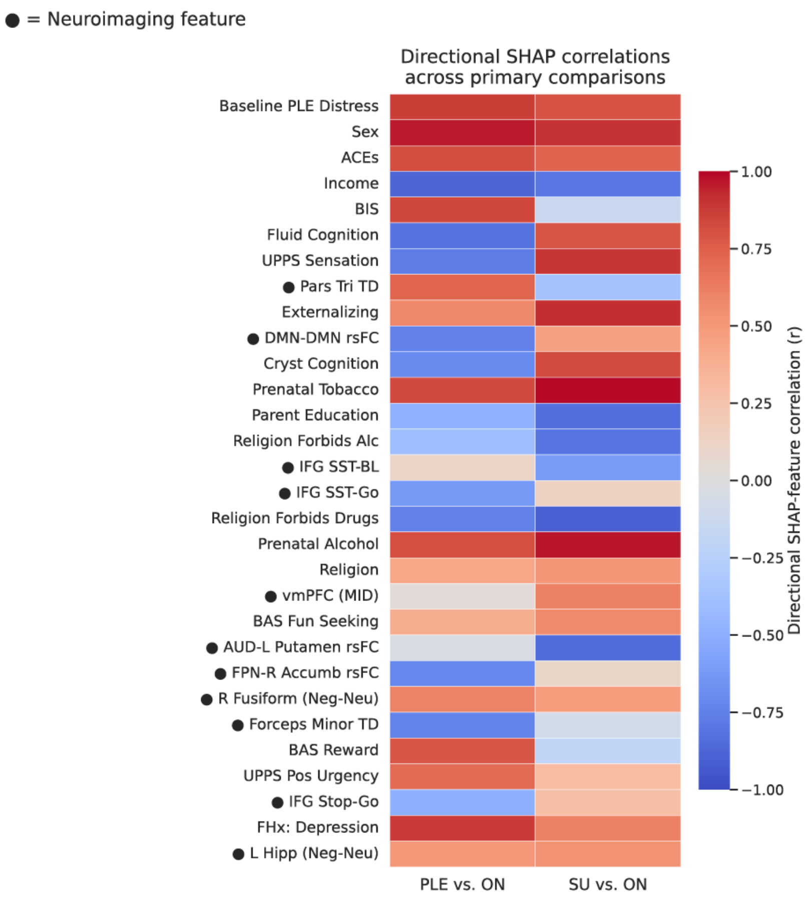


Directional feature importance for psychotic-like experiences (PLEs) and substance use (SU) derived from XGBoost SHAP analyses. A) Scatterplot showing directional associations between each feature and its SHAP value for SU (x-axis) and PLEs (y-axis), computed as the sign of the correlation between feature values and SHAP contributions on held-out test data, aggregated across cross-validation splits (see Supplementary Methods, Section 8). Displayed features comprise the union of the top 30 features ranked by combined PLE and SU mean absolute SHAP value and the top features identified across the three primary comparisons, ensuring that features contributing substantially to any single-outcome or co-occurrence model are represented. Each point represents a feature; triangles denote neuroimaging features and circles denote non-neuroimaging features. Distance from the origin reflects the strength of directional influence, with positive values indicating risk-increasing associations and negative values indicating protective associations. Dashed lines denote zero correlation, separating concordant and divergent effects across outcomes. Sex at birth was coded as binary; positive SHAP values indicate higher risk associated with female sex. Abbreviations: ACEs = Adverse Childhood Experiences; BIS = Behavioral Inhibition

System; UPPS = UPPS Impulsive Behavior Scale; Pars Tri ON = Transverse diffusivity (grey matter) of the IFG pars triangularis; DMN = Default Mode Network; rsFC = resting state functional connectivity; Cryst cognition = crystallized cognition; IFG = inferior frontal gyrus; SST = Stop Signal Task; IFG Stop-go: IFG activation during response inhibition (SST: inhibition vs. go); vmPFC = ventromedial prefrontal cortex; MID = Monetary Incentive Delay Task; BAS = Behavioral Activation System; AUD = Auditory network; Accumb = nucleus accumbens; FPN = frontoparietal network; R fusiform = right fusiform gyrus; Neg-Neu = the negative faces versus neutral faces contrast from the Emotional nBack Task; Forceps Minor TD = restricted total diffusion (white matter) of forceps minor; L Hipp = left hippocampus.

**Supplementary Figure 6:** SHAP Beeswarm Plots for Top Predictors by Comparison

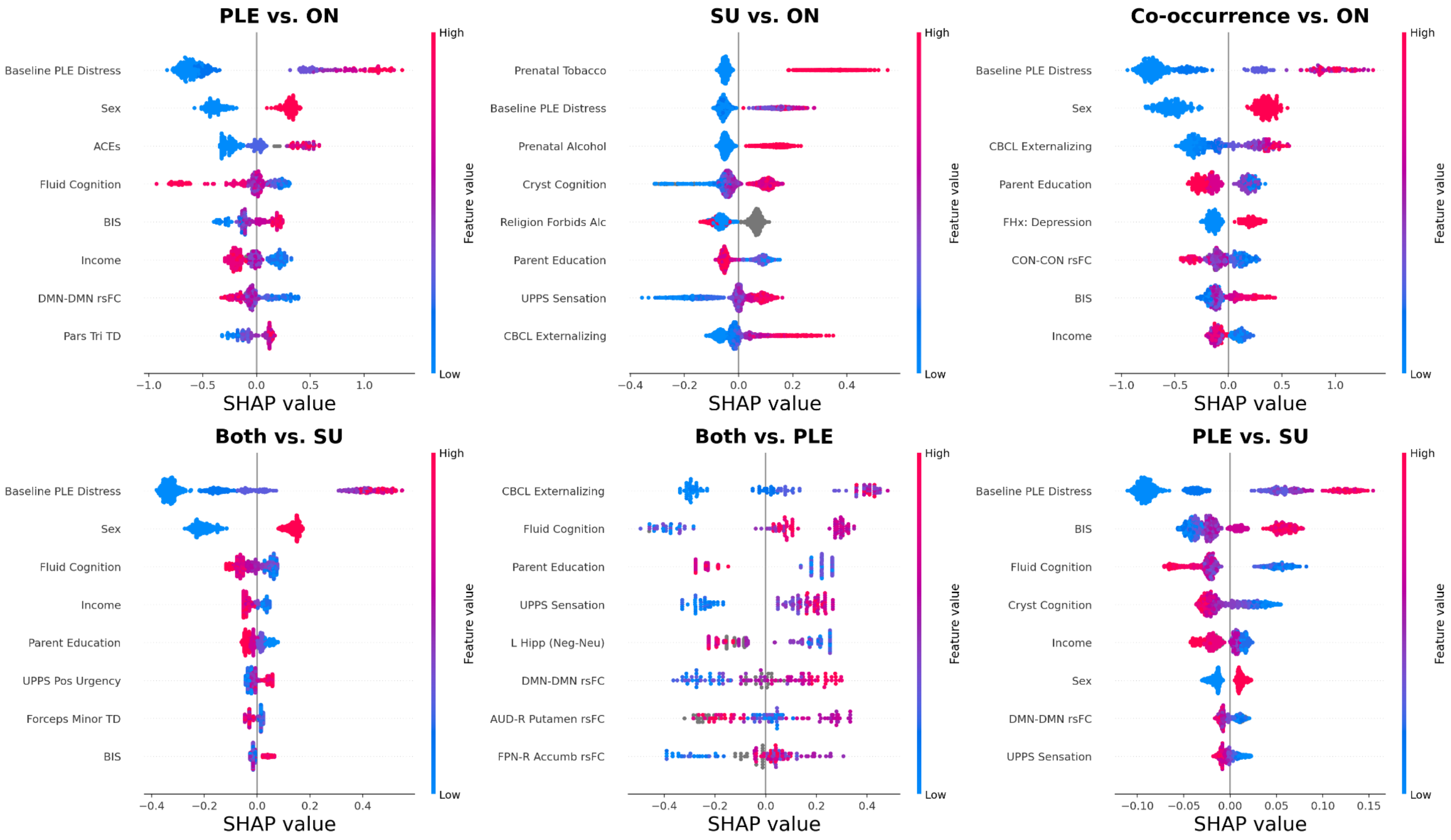


SHAP beeswarm plots showing the top 15 predictive features for each of the six binary comparisons, derived from the best-performing XGBoost cross-validation split using all features combined. Each point represents one test set observation. The x-axis reflects the SHAP value, the magnitude and direction of each feature's contribution to the model's prediction for that individual, and points are colored by the feature's raw value (red = high, blue = low). Features are ordered vertically by mean absolute SHAP value, with the most predictive features at the top. This figure illustrates both the relative importance of each feature and the direction of its effect on predicted outcome membership.

**Supplementary Figure 7:** SHAP Mean Absolute Value Bar Plots by Comparison

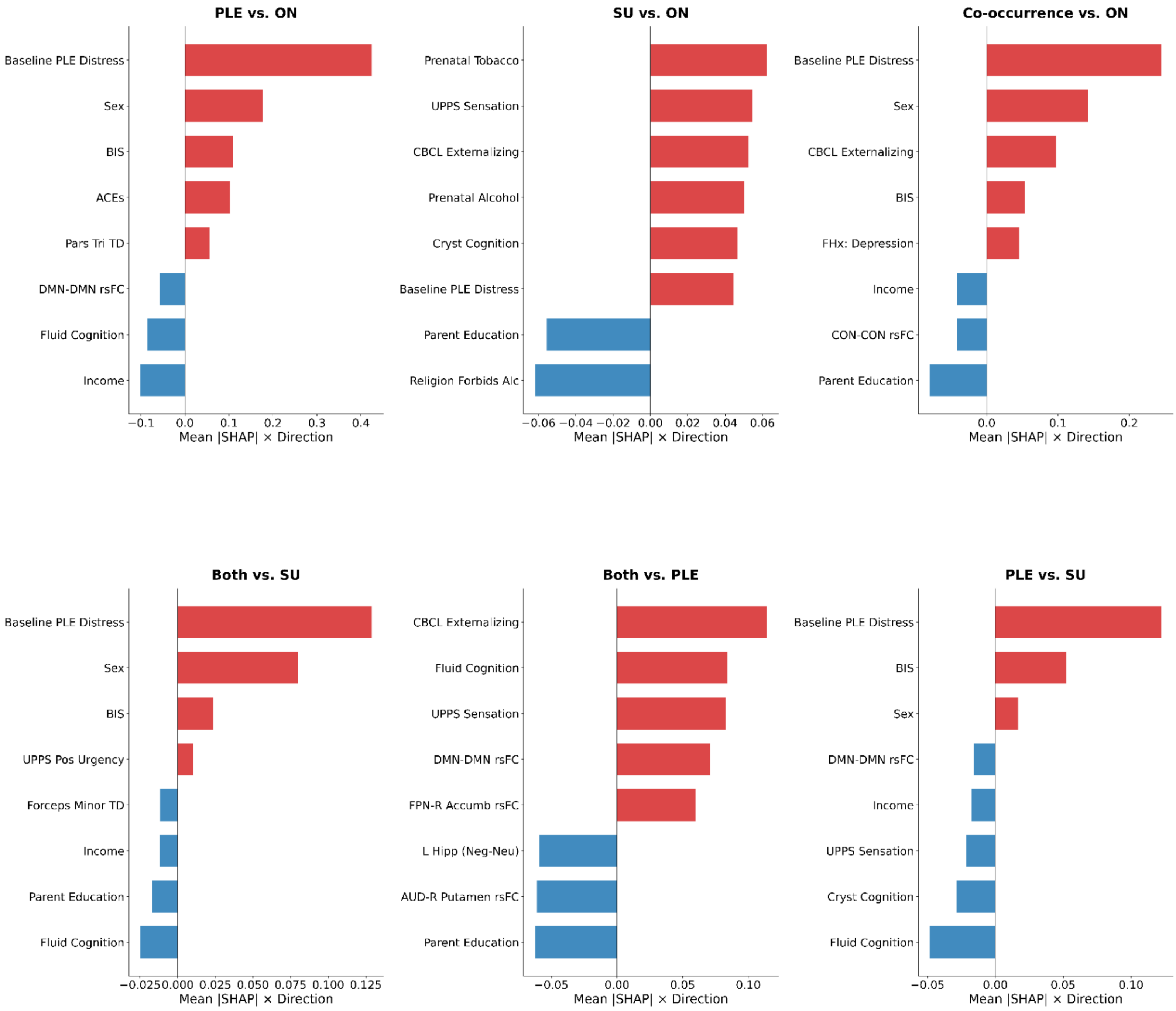

Directional SHAP feature importance for the top 8 predictors across all six binary comparisons, averaged across 5 cross-validation splits (XGBoost, all features). Bar length reflects mean absolute SHAP value and bar direction reflects the sign of the feature-SHAP Spearman correlation; that is, whether higher feature values increase (red, rightward) or decrease (blue, leftward) predicted probability of membership in the positive class. Features are ordered by signed SHAP value. This figure complements the beeswarm plots in Supplementary Figure 3 by providing a stable, averaged summary of feature directionality across splits rather than a single split snapshot.

**Supplementary Figure 8:** ROC Curves for Secondary Models

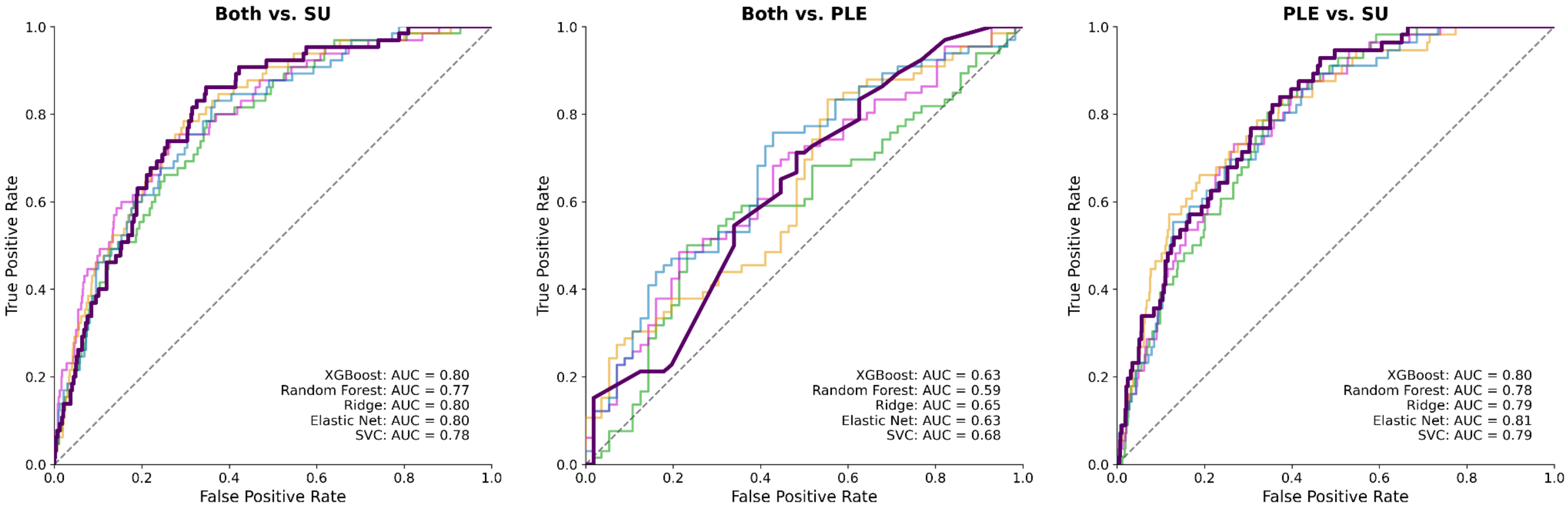


Receiver operating characteristic (ROC) curves for the three secondary binary comparisons (co-occurrence vs. SU-only, co-occurrence vs. PLE-only, and PLE-only vs. SU-only) across all five model types (XGBoost, Random Forest, Ridge, Elastic Net, SVC) using the full feature set. AUROC values reflect performance on the best-performing cross-validation split. The diagonal dashed line represents chance performance.

**Supplementary Figure 9:** Neuroimaging-only Permutation Tests

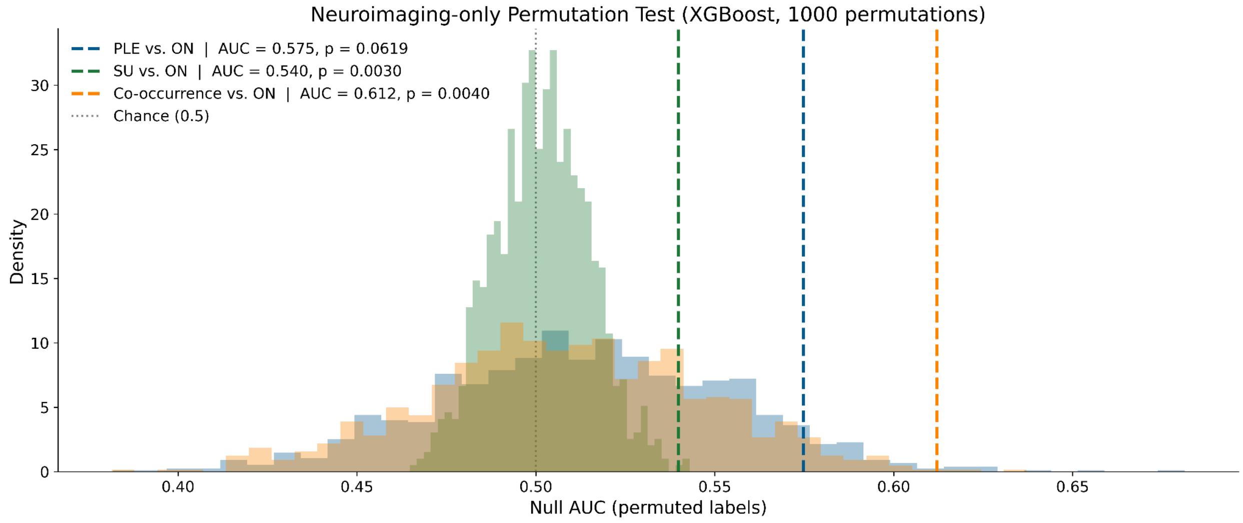


Permutation test results for neuroimaging-only models. Null distributions (1000 permutations each) from XGBoost classifiers trained on neuroimaging features only, for each primary outcome comparison: PLEs vs. ON (blue), SU vs. ON (green), and co-occurrence vs. ON (orange). Dashed vertical lines indicate observed mean test AUROCs across outer cross-validation splits. Permutation p-values were computed as the proportion of null AUROCs ≥ observed, with +1 smoothing in the numerator and denominator. SU and co-occurrence outcomes were significantly predicted by neuroimaging features alone ($p$'s $< 0.005$), while PLEs showed a marginal effect ($p = 0.062$).

**Supplementary Figure 10:** Toxicology positivity by modality

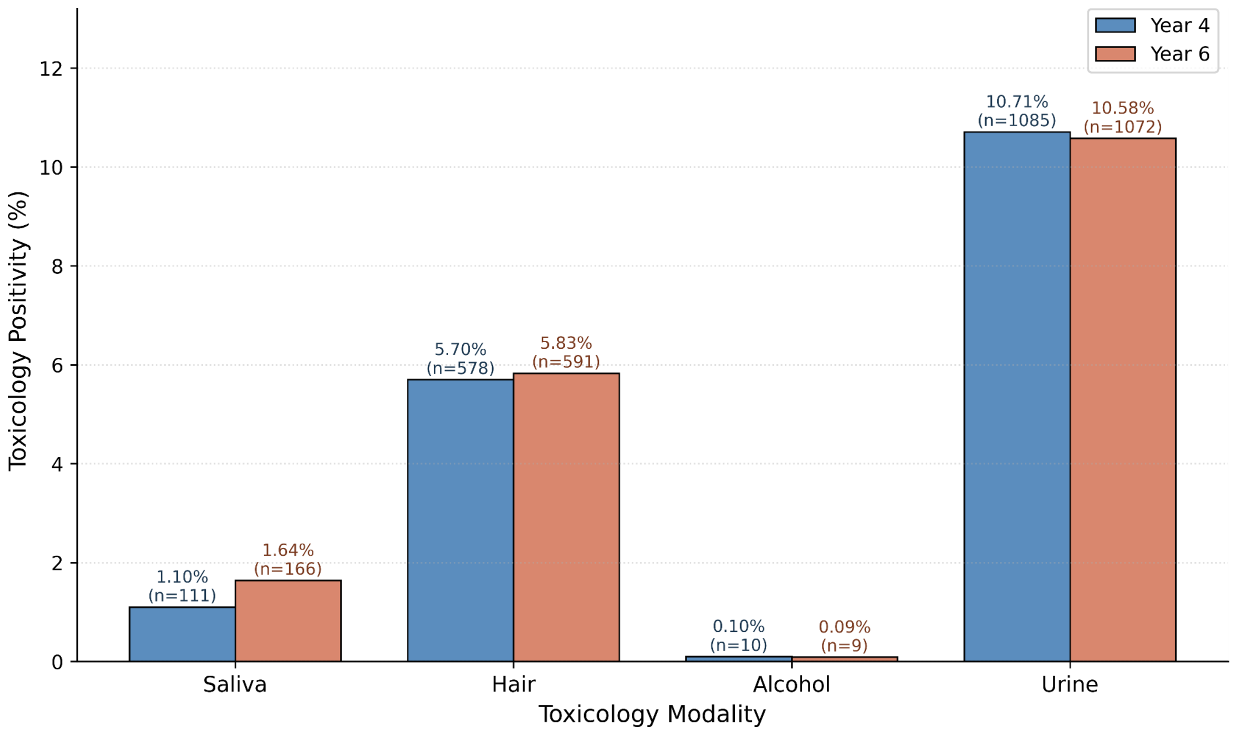


Toxicology positivity rates by modality at Year 4 and Year 6 follow-up, restricted to the analytic sample ($N$ = 10,134). Bars show the percentage of participants with at least one positive result on each toxicology modality, with raw counts annotated above each bar. Across both waves, urine drug screens yielded the highest positivity rate (~10.6–10.7%), followed by hair toxicology (~5.7–5.8%). Saliva (~1.1–1.6%) and alcohol-specific assays (~0.1%) yielded substantially lower positivity rates. Positivity rates were broadly stable across the two follow-up waves. Together, these toxicology modalities provided objective biospecimen evidence of substance exposure that complemented self-report, and any positive result across these modalities at Y4 or Y6 was used to define the substance use outcome (see Supplementary Methods 2).

**Supplementary Figure 11:** Feature Correlation Structure Within Sociodemographic Features

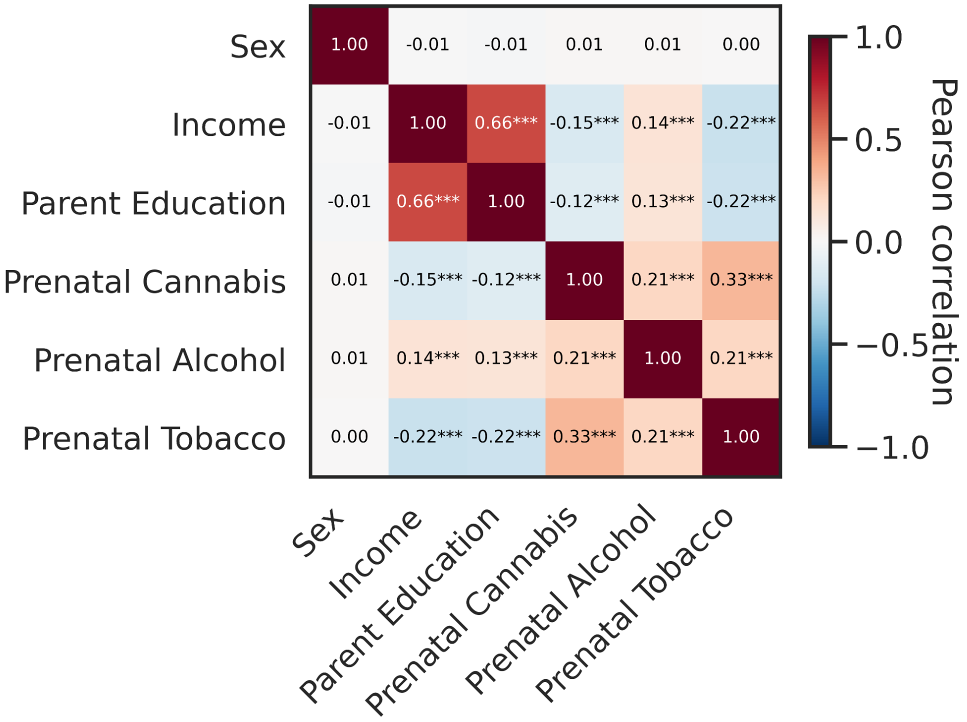


Pearson correlation matrix of sociodemographic features included in the baseline feature set. Correlation coefficients are displayed within each cell, with asterisks indicating statistical significance (**p* < .05, ***p* < .01, ****p* < .001). Income and parental education were strongly correlated ($r$ = .66), consistent with their shared variance as indicators of family socioeconomic status. Prenatal substance exposures showed moderate positive intercorrelations (*r*s = .21–.33), reflecting the known co-occurrence of prenatal substance use. Prenatal tobacco exposure showed the strongest correlations with lower household income ($r$ = -.22) and lower parental education ($r$ = -.22), while prenatal alcohol exposure was modestly positively correlated with both (*r*s = .13–.14). Sex was largely uncorrelated with the remaining features(rs ≤ .01). Given the large sample size (N = 10,134), statistical significance reflects sample size rather than effect magnitude; interpretation should focus on the magnitude of the correlation coefficients rather than on *p*-values alone.
